# Pontine pathology mediates common symptoms of blast-induced chronic mild traumatic brain injury

**DOI:** 10.1101/2023.01.26.23285066

**Authors:** James S. Meabon, Abigail G. Schindler, Daniel R. Murray, Elizabeth A. Colasurdo, Carl L. Sikkema, Joshua W. Rodriguez, Mohamed Omer, Marcella M. Cline, Aric F. Logsdon, Donna J. Cross, Todd L. Richards, Kole D. Meeker, Andrew Shutes-David, Mayumi Yagi, Daniel P. Perl, Desiree A. Marshall, C. Dirk Keene, William A. Banks, Ronald G. Thomas, Cory McEvoy, Adam Crabtree, Jake R. Powell, Jason P. Mihalik, Kathleen F. Pagulayan, Murray A. Raskind, Elaine R. Peskind, David G. Cook

## Abstract

Understanding how diffuse mild traumatic brain injuries can provoke common and persistent post-concussive symptoms (PCS), such as impaired sleep, is crucial to prevent and treat chronic disability and neurodegeneration. We mapped the spatially-resolved single cell landscape of diffuse mTBI pathology in a mouse model of blast exposure; identifying brainstem injuries predictive of later PCS. Repeated mTBI was necessary to establish chronic microglial activation and phagocytosis of myelin in the pontine reticular formation; where IL33 release by oligodendrocytes predicted microgliopathy. In postmortem brainstem tissues from patients with traumatic brain injury, chronic microglial activation and myelin phagocytosis was evident up to 20 years after diffuse mTBI caused by blast. In living patients with chronic blast mTBI, myelin injury in pontine projections mediated sleep disturbance and other PCS, with a dose dependent effect of mTBI number on sleep disturbance severity. These results support a mechanism for diffuse mTBIs to cause delayed persistent PCS.

## Main

Mild traumatic brain injuries (mTBIs), or concussions, account for ∼55 million annual global TBI cases.^1^ Although the causes of these injuries vary, persistent post-concussive symptoms (PCS) following mTBI—e.g., development of impairments in concentration, memory, mood, sensation, and sleep—are remarkably consistent.^2^ This consistency suggests that mTBIs share a common underlying neuropathology that may map to specific brain areas subserving these functions (e.g., brainstem nuclei). However, the mechanisms underlying these deficits are poorly understood, in part because mTBIs are undetectable on clinical neuroimaging.

Evidence suggests that TBIs may provoke the delayed development of deficits via neuroimmune axes that regulate the microstructural repair of the nervous system.^3,4^ This means that two critical determinants of long-term TBI outcomes are injury location and the subsequent cellular response. An improved anatomical understanding of specific TBI pathologies under conditions of diffuse injury, such as those caused by blast shock waves moving through the brain, could help predict the development of generalized functional or behavioral deficits following disease onset. Recent advances in high-dimensional spatially-resolved phenotyping coupled with precise anatomical brain mapping may help us understand how changes in the pathological microenvironment relate to persistent PCS following mTBI.

Even with minimal rotational head or body displacement, mTBI caused by exposure to explosive overpressures transmits a rapid force throughout the brain, diffusely injuring its tissues. The resulting pathology often differs from more kinetic forms of mTBI (e.g., caused by falls, assaults, etc.);^5^ however, blast-mTBIs often cause persistent PCS that are indistinguishable from other mTBIs.^6-11^ Therefore, we examined mTBIs associated with blast overpressure exposure in both military veterans and a mouse model of blast-mTBI with attenuated head movement.^12^ This approach avoids coup-contrecoup and “whiplash” injuries, while testing the hypothesis that blast-mTBIs provoke conserved post-concussive symptoms mediated by chronic pathological changes in brainstem nuclei.

## Results

### Brainstem nuclei are vulnerable to mTBI

Post-concussive symptoms typically relate to microscopic traumatic axonal injuries near small blood vessels^13,14^ in specific anatomical regions, such as cerebellum and corpus callosum,^13^ where vessel lesions can drive circuit dysfunction.^15^ Functional injury of microvascular tight junctions can be assessed by measuring the leakage of blood-borne molecules into the brain parenchyma. This method paradoxically shows that after TBI, disruption of the blood-brain barrier to the large molecule human albumin (66 kDa) is detectable prior to disruption to the smaller molecule sucrose (342 Da). However, disruption to sucrose persists longer than disruption to human albumin.^16^ We measured the functional integrity of the blood-brain barrier across brain regions 15 minutes to 3 days after a single blast-mTBI using blood-borne ^14^ C-sucrose and ^99m^ Tc-Albumin. Although a non-significant accumulation of sucrose and albumin was observed in the cortex during the acute phase (15 min after mTBI), significant leakages of both large and small molecules were detected in the brainstem, but not cortex (**Fig. 1a**). This indicates a regional vulnerability of the blood-brain barrier (BBB) to injury, with the brainstem’s BBB being more susceptible to injury after a single mTBI than the cortex. Injured neurons in areas of microvascular disruption can be identified by their uptake of blood-borne dyes that leak into the brain across the disrupted BBB, whereas adjacent uninjured neurons remain unlabeled.^17^ We used this method to determine the relative frequency of injured neurons across brain regions and white versus gray matter (**Fig. 1b, c**). Projection neurons, which are clearly distinguished from glia using this method (**Fig. 1d, e**), were labeled by extravasated fluorescent dye and occurred most frequently in the brainstem. In the brainstem, about 30% of mice showed evidence of neuronal injury within one hour and up to 70% of mice had observable neuronal injury within four hours of a single mTBI (**Fig. 1b**). Across sagittal brainstem sections, labeled cells occurred most frequently in white matter (**Fig. 1c**). Blood-borne labeling of injured brainstem neurons was visually more subtle than injured Purkinje cells of the inferior cerebellum (**Fig 1d**), a classic injury pattern of mTBI,^18^ likely owing to their expansive and dense arborizations.

**Figure 1.**
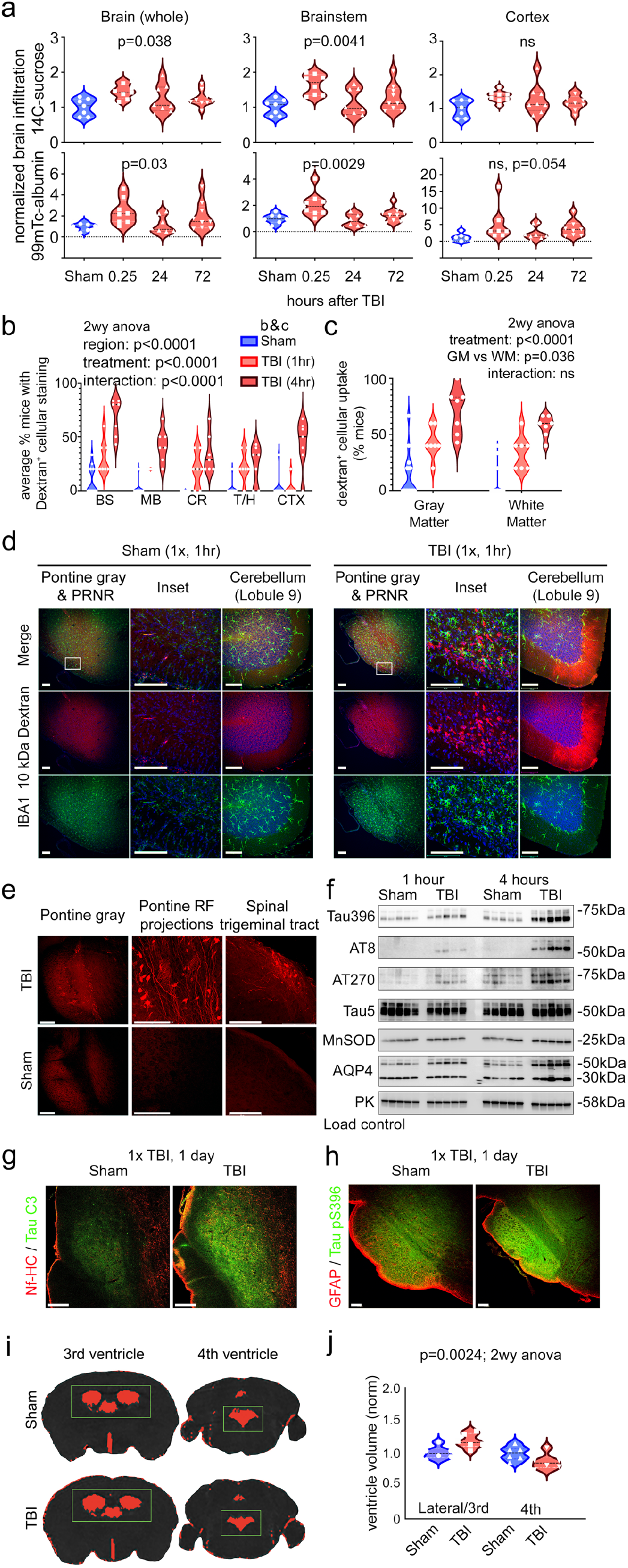
Pontine nuclei are vulnerableto a single blast mTBI. **a**, Time course and quantitation of blood-borne small (^14^ C-sucrose) and large (^99m^ Tc-albumin) molecule extravasation into brainstem,cortex, and whole brain. **b**, Time course and distribution of injured neurons labeled with a blood-borne fluorescent dye as observed using confocal microscopy. **c**,Injured cell distributions viewed across white vs. gray matter. (**b**) and (**c**) show the average number of animals per experiment with labeled cells (mice, 1 hr: N=35,35; 4 hr: N=40,40; average ∼5 mice per group per cohort; 7 cohorts). **d**, Representative micrographs of injured, dye-labeled neurons and accompanying microglial response 1 hr following TBI. Staining of injured pontine cells is subtle compared to cerebellar Purkinje cells. PRNR, rostral pontine reticular nucleus. **e**, Example micrographs of injured locations often observed. RF, reticular formation. **f**, Western blots document increasing tauopathy (Tau396, AT8, AT270; axoninjury markers) and gliovascular injury (aqp4) during the 1-4 hr acute inflammatory phase after a single diffuse mTBI (5 mice/group). Tau5, total tau; PK, pyruvate kinase (load control). **g** and **h**,Confocal micrographs of (**g**) caspase-cleaved Tau (Tau C3) and (**h**)pathologically associated tau hyperphosphorylation (pS396) in the pons1 day after TBI. **i**, Representative T2 MRI of mice 24 hrs after a single TBI or shamprocedure. Red color indicates pixels that exceed the threshold value (representative of CSF) and green is the region-of-interest(ROI) bounding box. **j**, Quantification of ventricle volumes (6 mice/group). One-way ANOVA (**a**), two-way ANOVA (**c, j**),and mixed effects model (**b**). Scale bars are 100µm.

Injured brainstem neurons and areas of BBB damage coincided with local changes in microglial morphology that are consistent with activation and phagocytosis of extravasated materials (**Fig. 1d**). Diffuse axonal and neuronal injuries were primarily observed in the pontine reticular formation (RF), one of several brainstem regions controlling sleep behavior (**Fig. 1e**). Localized cellular injuries were observed laterally toward the brainstem surface, with frequent prominent injuries along the ventral caudal surface, spinal trigeminal tract, and dorsal columns and was independently identified by unguided investigators (AFL, KDM) blinded to the study groups.

The development of brainstem tauopathy, indicating axon injury, paralleled increased expression of aquaporin-4 (AQP4), which regulates the brain-wide convection of water. A compensatory increase in the expression of magnesium superoxide dismutase (MnSOD), a mitochondrial redox enzyme responsive to TBI-related cellular injury,^12^ paralleled the increase in the fluorescent labeling of injured neurons (**Fig. 1f**). Tauopathy in the pontine gray, medial lemniscus, and RF persisted at least 24 hours after injury (**Fig. 1g, h**).

Cerebral edema often follows both clinical and experimental mTBI despite unremarkable neuroimaging results. We postulated that the prominence of diffuse injury from the foramen magnum to the 4^th^ ventricle and the related changes in the AQP4 water transporter would be accompanied by tissue swelling. Such swelling, we posited, would result in reduced 4^th^ ventricle volume per T2 MRI and be associated with secondary cerebral edema, yielding enlargement of the 3^rd^ and lateral cerebral ventricles. In group-blinded analyses of mice assessed 24 hours after a single blast-mTBI, we found that mTBI induced significant enlargement of the lateral and 3^rd^ ventricular volume while reducing the 4^th^ ventricular volume (**Fig. 1i, j**). These results indicate that a single blast-mTBI can cause axonal injury and inflammation of the brainstem in the absence of gross hemorrhage. Such injuries may contribute to more expansive secondary injuries related to edema throughout the brain.^19^

### mTBI establishes persistent brainstem microgliopathy

Repetitive mTBI is an environmental risk factor for chronic neurodegenerative phenotypes.^20-22^ Genetic risk factors for neurodegeneration often map to myeloid cells, including peripheral blood monocytes and brain resident microglia.^23^ However, our understanding of how disease-associated immune phenotypes relate to TBI pathologies is incomplete. To determine the relationship between myeloid phenotypes and the neuropathological landscape of the tissue microenvironment after repeated mTBI, we first tested the *a priori* hypothesis that functional alterations in brain myeloid cells would be a persisting and sensitive indicator of regional injury, with the greatest changes occurring in the brainstem as seen during acute injury. To evaluate this hypothesis, we used a RiboTag strategy to assess the active translatome specifically in brain myeloid cells from mice with repeated TBI or sham exposure (1 per day for 3 days), avoiding the trauma-induced changes in gene signatures evoked by *ex vivo* dissection of live tissue and cell sorting that are known to occur.^24,25^ By using a tamoxifen-inducible CX3CR1-creER mouse line shown to be superior in achieving brain macrophage specificity with the exclusion of neuron-derived contaminating mRNA,^24^ we employed a discovery approach followed by validation in an additional secondary cohort to identify candidate genes differentially expressed across brainstem, cerebellum, and cortical tissues in mice one month after repetitive mTBI (3x; 1 per day for 3 days), as compared with gene levels in tamoxifen-induced CX3CR1-creER control mice receiving an equal number of sham-TBI treatments (**Fig. 2a, b, c**). Brainstem tissues showed more differentially expressed myeloid genes than cerebellar or cortical tissues consistent with acute injury patterns being concentrated in the brainstem.

**Figure 2.**
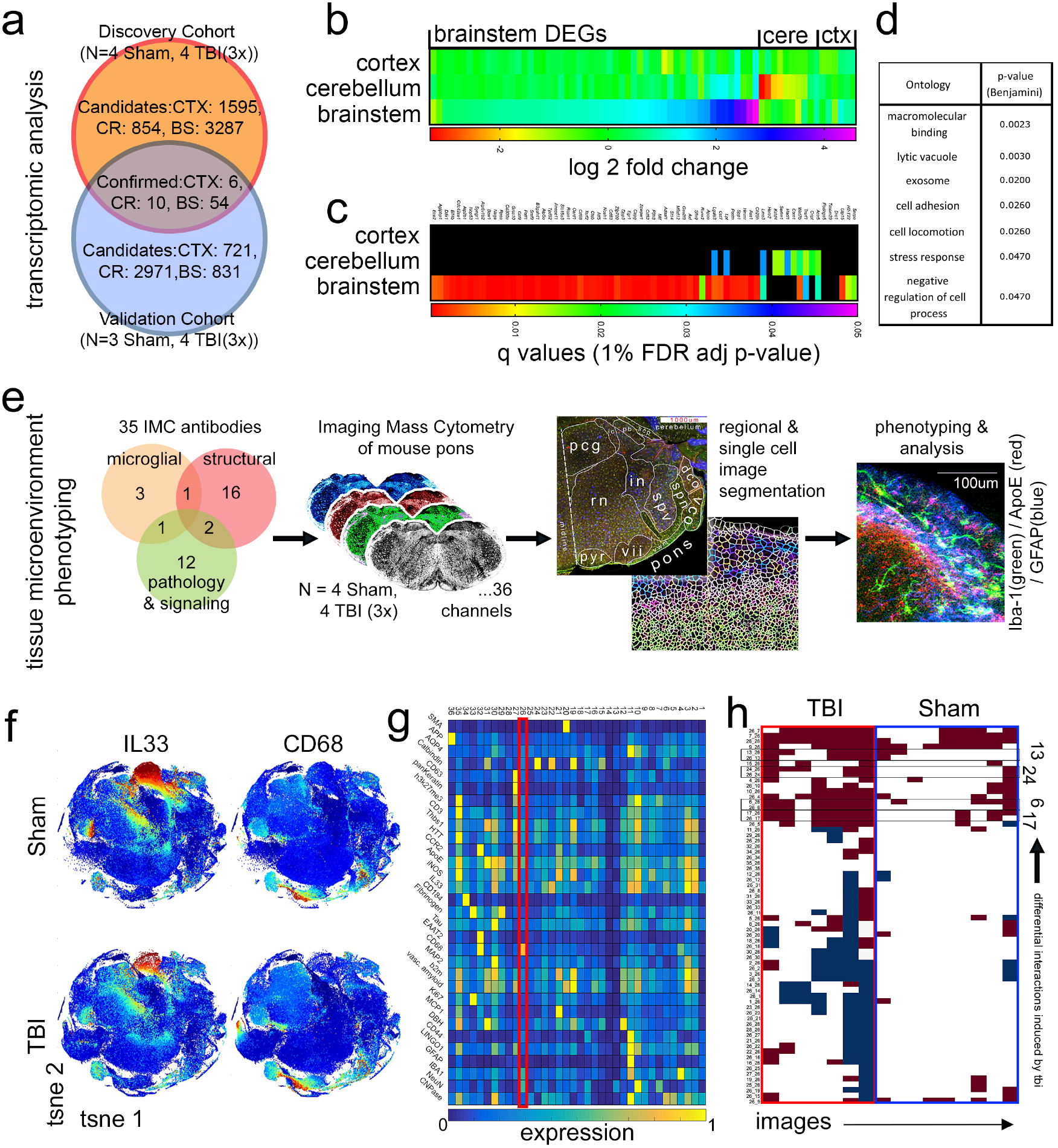
mTBI induces persistent pontine microgliopathy. **a**, Discovery and validation strategy using 3-month-old male Cx3cr1^CreER^ :Rpl22^HA^ RiboTag mice to identify genes actively transcribed and differentially expressed in myeloid cells one month after repeated (3x) mTBI as compared to sham control mice. **b**, Heatmaps of candidate differentially expressed genes (DEGs) identified by RNA sequencing across brain regions. **c**, False discovery rate-adjusted p-values for candidate DEGs. **d**, Gene Ontology terms significantly associated with brainstem DEGs. Benjamini adjusted p-values. **e**, Imaging mass cytometry (IMC) workflow for detecting and phenotyping changes in brain tissue microenvironments after injury. **f**, T-SNE-based global phenotyping of single-cell image data. **g**, Phenograph analysis with unbiased identification of disease associated microglia (DAMs) (cluster 26, red box; CCR2^-^, Iba-1^+^, CD68^hi^, ApoE^hi^). **h**, Neighborhood analysis identifying changes in the frequency of associations between DAMs (Phenograph cluster 26) and phenograph-defined tissue microenvironments.

We then performed gene ontology (GO) term analysis. Given that significantly fewer genes were confirmed in the cortex and cerebellum, only differentially expressed genes confirmed in brainstem tissue were used (**Fig. 2d**). In agreement with previous reports, blast-mTBI–induced geneset enrichments in brainstem myeloid cells included programs for macromolecular binding (e.g., *APOE*), lytic vacuole (e.g., *CD68*), exosome, cell adhesion and locomotion.^23^

Given that microglial properties shift in response to the conditions of their tissue microenvironments, we developed an imaging mass cytometry (IMC) workflow to visualize how the relations between microglial phenotypes and their tissue microenvironments changed in response to TBI (**Fig. 2e**). To do this, we developed a panel of 35 metal-labeled antibodies specific to murine TBI histology, including cellular, pathological, and signaling markers (**Table S2**). Antibody-stained tissue slides were then laser-ablated and the subsequent signal was detected with time-of-flight mass spectrometry, producing a 2-dimensional, rasterized representation of metal abundances analogous to a highly-multiplexed microscope image. Following *in silico* image segmentation into single cells in CellProfiler, analyses were conducted in HistoCAT and R. We identified 256,295 cells from images of 8 coronal pontine hemisections that were taken from randomly selected study mice and that provide tissue representation of ventral pontine and dorsal structures near the 4^th^ ventricle (**Fig. 2e)**.

Phenotyping based on nonlinear dimensionality reduction was implemented using Barnes-Hut t-distributed stochastic neighbor embedding (t-SNE) (**Fig. 2f; Fig. S1**). To define the prominent microenvironmental phenotypes of single cells in the injured brain, we used unbiased phenograph-based clustering to yield 36 distinct clusters for further analysis (**Fig. 2g**). This method differentiated both unreactive microglia (cluster 4; Iba^+^, CCR2^-^, CD68^lo^, ApoE^lo^) and a disease-associated microglial (DAM) phenotype (cluster 26; Iba^+^, CCR2^-^, CD63^+^, CD68^hi^, ApoE^hi^) consistent with single-cell RNA sequencing analyses of Alzheimer’s disease brain and neurodegenerative models,^23,26,27^ in addition to several non-microglial phenotypes representing neuronal, vascular, glial, and peripheral immune cells. Intercellular associations, as measured by neighborhood analysis, increased between spatially resolved disease-associated microglia with IL33^+^ myelin (clusters 6 and 17), NeuN^+^ neurons (cluster 13) and HTT^+^ neurons (cluster 24) in brain-injured mice whereas similar associations did not change in control mice (**Fig. 2h**). Taken together, these findings suggest that repetitive mTBI provokes a persisting pontine microgliopathy that is associated with slowly resolving neuronal and IL33^+^ myelin injury, which can be spatially resolved and quantified using unbiased computational means.

### Myelin injury precedes sleep deficits

Delayed-onset PCS are thought to reflect progressive changes in cellular phenotypes prior to functional and structural brain changes. We used a data-driven approach to agnostically identify specific regions of persisting brainstem pathology that could cause persistent PCS. More specifically, we gated on disease-associated microglia in IMC images and measured their average frequency distributions (i.e., DAM/total microglia) per subject across exclusive brain-region image masks of whole pontine hemisections. We then used the results to inform later hypothesis testing on the relations between pathology, chronic symptoms, and the known functions of the brain regions. Representative IMC images of pontine disease-associated microglia in blast-mTBI and sham control mice are shown in **Figure 3a**. TBI increased the frequencies of disease-associated microglia in several areas with post-hoc FDR-adjusted analyses identifying notable increases in the reticular formation (rf; t[62]=3.366, q=0.0019), spinal trigeminal nerve tract (spn; t[62]=3.066, q=0.0034), ventral white matter (vwm; t[62]=3.317, q=0.0019), and ventral cochlear nucleus (vco; t[62]=6.448, q<0.0001) (**Fig. 3b**). Since blast-mTBI induces the association of DAM with myelin expressing the alarmin molecule IL33 (**Fig. 2i**), we examined the expression of IL33 among all 44,023 spatially resolved single CNPase^+^ /IL33^+^ oligodendrocytes distributed across these regions. Post-mTBI oligodendrocytes of several pontine regions showed significant loss of the alarmin IL33 (vii: t[42]=1.98, q=0.05; rf: t[42]=3.225, q=0.0062; in: t[42]=3.406, q=0.0062; spv: t[42]=3.108, q=0.0062; spn: t[42]=2.782, q=0.0118; pyr: t[42]=2.3, q=0.0278; pcg: t[42]=2.711, q=0.0118; **Fig. 3c**), through its apparent release from nuclear stores into the surrounding microenvironment (**Fig. 3d**). Microglia, which express IL33R mRNA (**Fig. 3e**), accumulate in a predictive linear manner driven by IL33 levels (**Fig. 3f**).

**Figure 3.**
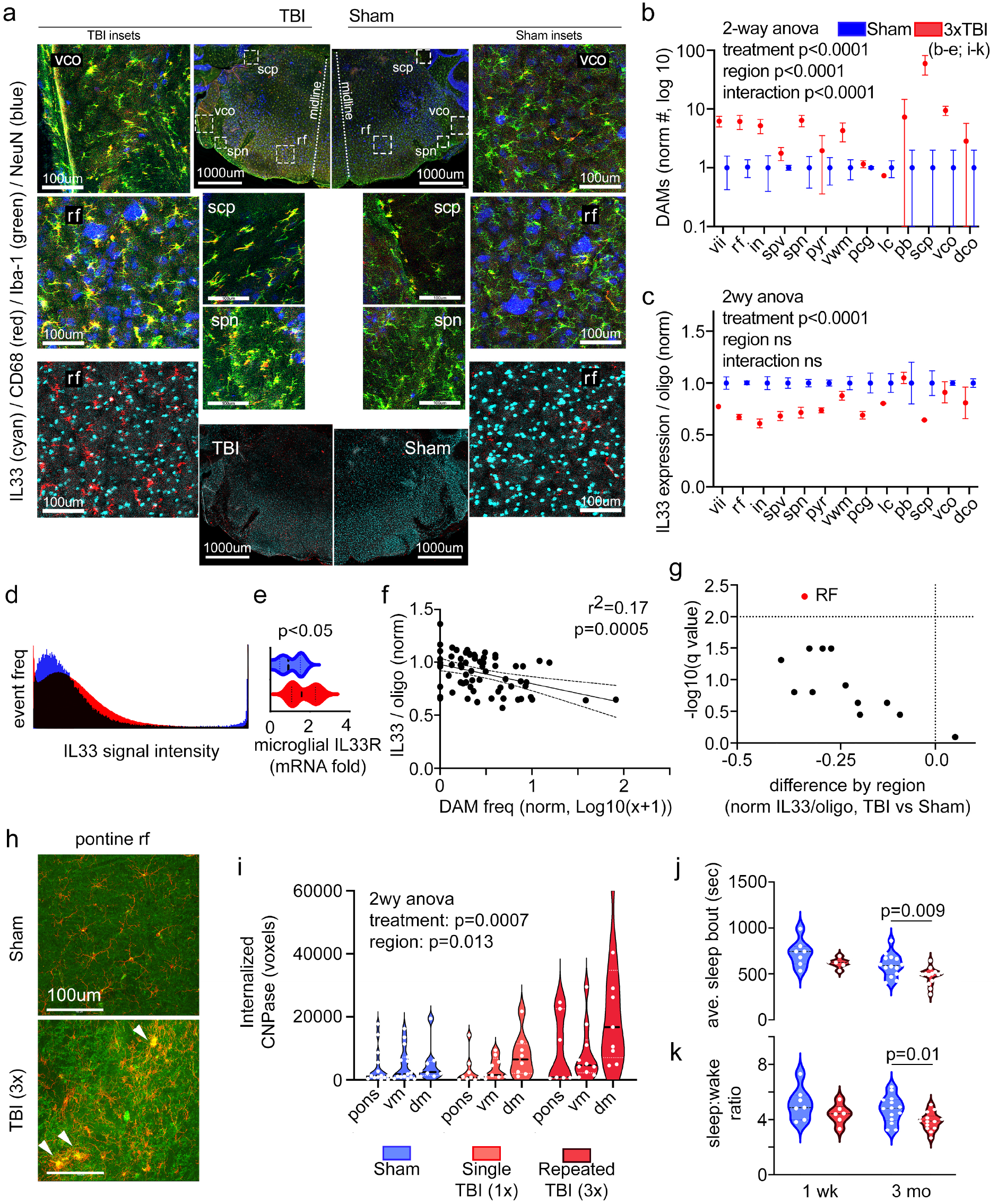
mTBI-induced IL33 release by oligodendrocytes is associated with microglial-mediated myelin remodeling. **a**, Representative IMC images of pontine hemisections from repeated TBI and sham mice. **b**, Average frequency of DAM per subject detected in IMC across pontine regions. **c**, Average expression of oligodendrocyte IL33 per mouse measured across pontine regions. **d**, Redistribution of nuclear oligodendroglial IL33 into the surrounding parenchymal microenvironment measured as a frequency histogram of IL33 signal intensity across pixels from insets in (**a**). **e**, Quantitation of microglial-specific IL33R transcript determined from microglial RiboTag mice (N=7 sham, 8 TBI (3x)). **f**, Release of oligodendroglial IL33 predicts the accumulation of DAMs. DAM frequency significantly increased with loss of oligodendroglial IL33 among all pontine regions. Linear regression model. **g**, Volcano plot of q values (FDR-adjusted p-values) identifying robust myelin injury in the pontine RF, as measured by IL33 depletion from oligodendrocytes. **h**, Example of maximum intensity projection images from 3D confocal micrographs of microglia (red, Iba1) and myelin CNPase (green). Arrowheads denote microglial nodules consuming myelin. **i**, Violin plots show the total amount of CNPase internalized by microglial cells in each 50µm x 632µm^2^ 3D confocal image. **j**, Length of average sleep bouts and (**k**) sleep-to-wake ratios at 1 week and 3 months after repeated TBI. Red represents TBI data; blue represents sham control data. Data are a distribution of measured means from individual mice. Regions in **(b**,**c**,**g)** correspond to map in **Fig. 2e**. dm, dorsal medulla; rf, pontine reticular formation; scp, superior cerebellar peduncle; spn, spinal trigeminal nerve tract; vco, ventral cochlear nucleus; vm, ventral medulla.

Next, we used a false-discovery rate-adjusted method to focus our analysis on the pontine structure(s) displaying the most consistently altered IL33 expression following TBI. We found highly significant mean IL33 reductions per oligodendrocyte for each blast and sham mouse in the RF indicating robust and persisting white matter injury (**Fig. 3g**). Disease-associated microglia heavily express the lytic vacuole protein CD68, which is known to facilitate phagocytotic clearance of diseased tissue, including myelin.^17,28^ CNPase is an integral myelin protein required for axon paranode maintenance^29^ in that its axoglial decoupling is associated with progressive axonal degeneration^30^ and phagocytosis by microglia during dysmyelination^29^ and Wallerian degeneration.^31^ Because IMC is limited to 2D analyses, we used 3D confocal microscopy of sagittal brainstem sections near midline to measure how microglial internalization of CNPase relates to both injury location across brainstem structures and the number of blast-mTBIs administered. Phagocytosis of CNPase by both single-cell and clustered microglia was observed in pons and medulla, with greater apparent co-localization observed in microglial nodules than single microglia (**Fig. 3h, arrowheads**). One month after single or repeated (3x) blast-mTBI, myelin consumption was measured as the number of CNPase-positive voxels inside Iba-1-stained microglial volumes. Myelin consumption by microglia increased in relation to the number of mTBIs (F(2,85)=7.88, p=0.0007) and anatomical location (F(2,85)=4.536, p=0.0134)(**Fig. 3i**), with Tukey’s post-hoc analyses determining these effects were driven by repeated blast-mTBI (sham vs 1x.: 444 mean dif. (−5150 to 6037, 95%CI), q=ns; sham vs. 3x: - 8336 mean dif. (−13961 to -2711, 95%CI), q=0.0019).

Sleep impairment following TBI occurs in 30–65% of those with persistent PCS.^32,33^ Interestingly, insomnia, which is characterized by difficulty initiating and maintaining sleep, is more common with less severe TBIs and blast-related mTBIs.^34-36^ To measure changes in post-traumatic sleep that are caused by blast-mTBI and that may contribute to the development of later sleep impairment, we used a noninvasive sleep monitoring system to record changes in the stereotypical sleep patterns of mice.^37,38^ When measured seven days after blast-mTBI, the sleep bout duration and diurnal sleep-to-wake ratio of blast-mTBI mice was normal compared to sham control mice (**Fig. 3j, k**) and was consistent with previous reports of TBI mice one week after injury.^39^ However, following a three-month period of recovery from repeated blast-mTBI, injured mice developed a statistically significant increase in sleep fragmentation characterized by a reduced average sleep bout duration and lower diurnal sleep:wake ratio (**Fig. 3j, k**) that was similar to clinical insomnia in humans. Thus, the development of sleep impairment occurs after the onset of microglial-associated pontine myelinopathy.

### Myelin injury mediates sleep injury

We hypothesized a similar pathology would exist in the brainstem of military veterans with blast-mTBI. In a neuropathological analysis of autopsy subjects with (n=4) and without (n=4) blast-mTBI, phagocytic microglial nodules that highly express CD68 (i.e., microgliopathy; **Fig. 4a**) were often seen with internalized myelin (i.e., CNPase; **Fig. 4b**). Microglia nodules were identified in the brainstem and cerebellar white matter tracts of subjects up to 20 years after their last reported TBI. By observing IMC images stained with 6 antibodies specific for human protein targets, we confirmed that human microglial nodules were phagocytic and transcriptionally active (i.e., CD68^+^, negative for histone 3 lysine 27 trimethylation) in contrast to the adjacent, uninvolved, normal-appearing microglia (**Fig. 4c**).

**Figure 4.**
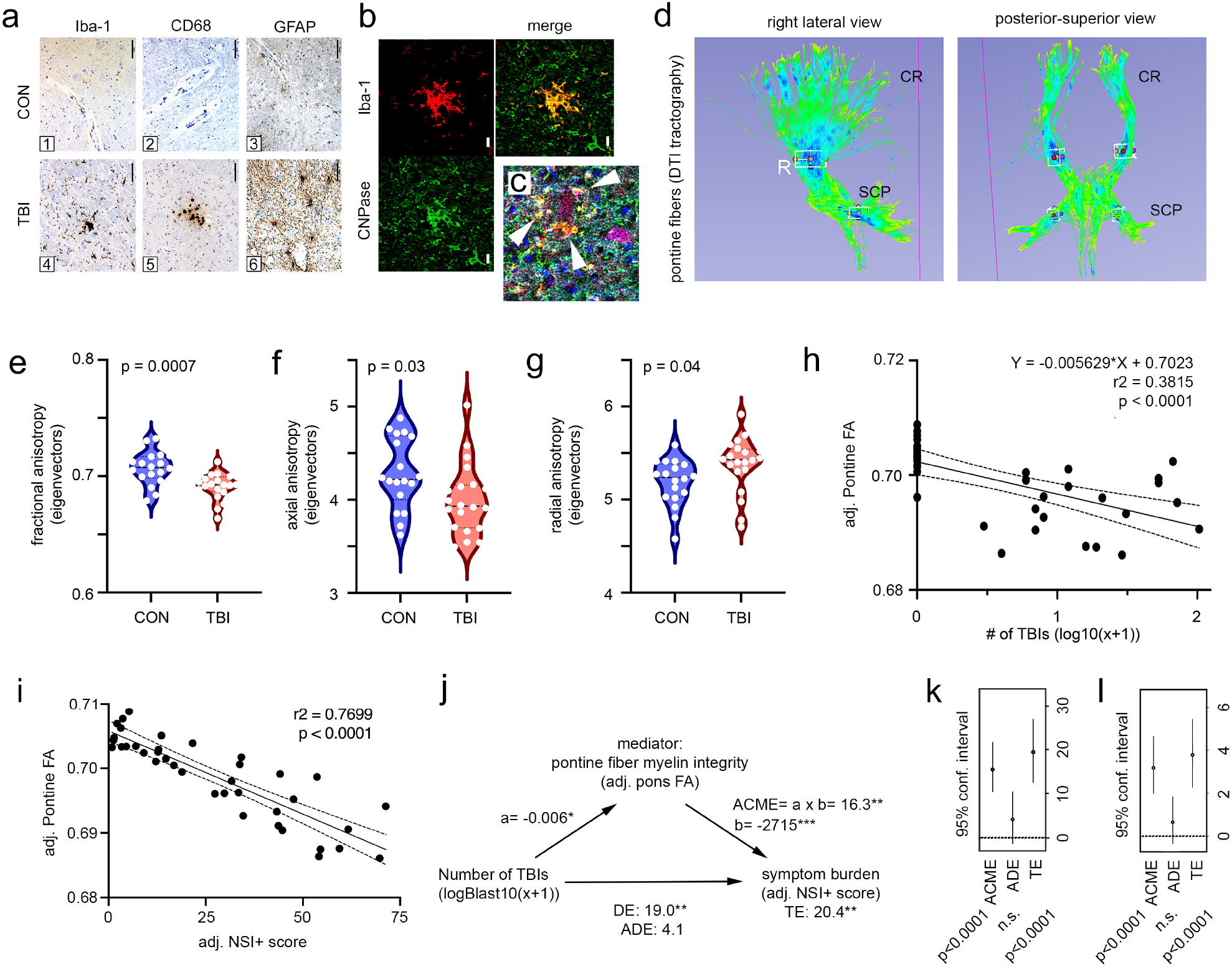
Pontine myelin injury mediates chronic blast-mTBI symptoms. **a**, Representative examples of brainstem glia in blast-mTBI and control subjects observed in three separate clinical neuropathology labs. **b**, Example confocal micrograph of clinical microglial nodule (red) with apparent internalization of myelin CNPase (green). **c**, IMC of pontine DAM (arrowheads) phagocytosing a neuron as seen in a clinical TBI subject. Purple, NeuN; cyan, CNPase; green, Iba-1; red, CD68; white, GFAP; blue, H3K27Me3. **d**, DTI tractography generated from pontine seed points. Boxes indicate the approximate locations of volumes of interest (VOIs) used for clinical DTI measures of major pontine projection tracts. Seed points were selected based on the acute pontine injury pattern observed in our animal study. CR, corona radiata; SCP, superior cerebellar peduncle. **e–h**, Quantification of chronic myelin injury is averaged across VOIs in living subjects. **e**, DTI fractional anisotropy (FA) is reduced in TBI subjects compared to control subjects; with groupwise changes in (**f**) axial and (**g**) radial anisotropy. **h**, Cumulative TBI-dose dependent decreases in pontine FA values adjusted for age and PTSD severity using linear regression modeling. **i**, Path diagram for causal mediation modelled with multivariable linear regression. ACME: adjusted causal mediation effect estimate, DE: direct effect estimate, ADE: adjusted direct effect estimate, TE total effect estimate. *p<0.05, **p<0.01, ***p<0.001. **j**, ACMEs for NSI+ total score related to blast-mTBI as mediated by reductions in mean pontine fiber FA (adjusted for age and PTSD). **k**, Results of bootstrap validation of mediation path illustrated in **(j). l**, Boot-strap validated mediation analysis of blast-induced pontine fiber injury to cause sleep injury using PTSD and age-adjusted PSQI total scores. Two-tailed t-tests in (**e**-**g**). Scale bars are (**e**) 100µm and (**b, i**) 10µm.

Insidious myelinopathy mediates several clinical diseases including multiple sclerosis, Parkinson’s disease, and stroke and may contribute to the earliest occurring changes in age-related cognitive decline and disability following TBI.^40^ Our findings in mice indicate that diffuse axonal injury in the brainstem initiates latent white matter pathology that may drive the development of a spectrum of delayed-onset behavioral impairments related to mTBI. Diverse cognitive, behavioral, and somatic PCS occur in 77–97% of persons with repetitive TBI.^35,41^ These persistent PCS develop over hours to months following injury^42^ and may become chronic, lasting years.^43^

To determine whether brainstem white matter changes caused by diffuse axonal injury are associated with neurobehavioral symptoms in humans with blast-mTBI, we used magnetic resonance diffusion tensor imaging tractography (DTI) in living US veterans with repetitive blast-mTBI (n=20) and veterans deployed to the Iraq/Afghanistan war zones with no lifetime history of TBI (n=19). Subject demographics are provided in **Supplemental Table 3**. Guided by our animal model (**Fig. 3**), we reconstructed fiber tracts using the entire human pons as the seed region to avoid placement bias or error (**Fig. 4d**). Consistent with our observation of TBI-related myelin and neuronal injury in the pons of mice, TBI was associated with significant myelin injury of pontine fibers in veterans with mTBI via measures of DTI fractional, axial, and radial anisotropy (**Fig. 4e-g**). We next evaluated a possible dose-response between the number of blast-mTBIs and changes in pontine fractional anisotropy (FA) while controlling for the potential confounding variables of age and posttraumatic stress disorder (PTSD) symptom severity. Reductions in adjusted pons FA were associated with increasing numbers of self-reported blast-mTBIs (linear regression, *p*<0.0001; **Fig. 4h**).

Diverse functions are controlled by highly conserved brainstem circuits, including functions related to the common symptoms following mTBI: sleep structure, emotional state, and sensorimotor integration. To learn whether the development of these symptoms may be mediated by changes in pontine white matter, we first evaluated the linear relation between age- and PTSD symptom severity-adjusted changes in pontine FA in all subjects and self-reported severity of persistent PCS related to mTBI as measured by the total score on an augmented Neurobehavioral Symptom Inventory (NSI+) questionnaire. Subjects rated symptom severities for dizziness, imbalance, dyscoordination; headache, nausea, vision, sensitivity to light; hearing, ringing in the ears, sensitivity to noise; numbness/tingling, changes in taste/smell; changes in appetite; changes in concentration, forgetfulness, decision-making difficulty, slowed thinking; fatigue, sleep impairment; anxiety, depression; irritability, poor frustration tolerance, getting into fights, disinhibition, mood swings; apathy/withdrawal; and slowness in speech ranging from 0 to 4 (not present, mild, moderate, moderately severe, severe) and symptom scores were summed (NSI+ total score). Using linear regression modeling, the relationship was highly significant (*p*<0.0001), indicating that greater total persistent PCS burden was associated with greater reductions in pontine FA (**Fig. 4i**). The number of lifetime mTBIs (log-10(x+1) transformed) similarly correlated with age- and PTSD symptom severity-adjusted measures of NSI+ total score (Spearman’s *r* = 0.630, 0.5024 to 0.7308 95% CI; *p*=2.7 × 10^−14^) indicating that total persistent PCS burden increases with the number of repeated mTBIs caused by blast exposure.

Lastly, we used statistical mediation analysis to evaluate the potential causal relationship between pontine white matter injury and persistent PCS burden. Using both pontine FA and NSI+ total scores adjusted for age and PTSD symptom severity, the regression coefficient between the number of blast-mTBIs and persistent PCS burden (i.e., the direct effect, DE) and the regression coefficient between pontine FA values and symptom burden were both significant (**Fig. 4j; Fig. S6a**). The indirect effect was (−0.006)*(−2715.00) = 16.3 (*p*<0.01). A validation analysis using nonparametric bootstrapping procedures determined that the effect size 95% confidence interval ranged from 9.460 to 23.05 (**Fig. 4k**). The validated adjusted causal mediation was statistically significant (*p*=2 × 10^−16^), supporting the conclusion that diverse persistent PCS following repetitive blast-mTBI are at least partially caused by pontine myelin injury. An identical mediation analysis determined a statistically significant relation between pontine FA and sleep impairment as measured by the Pittsburgh Sleep Quality Index (PSQI) total score similarly adjusted for age and PTSD symptom severity. Using a bootstrap-validated mediation analysis, both the adjusted causal mediation effect (ACME) and total effect (TE) were significant (ACME: 3.2 (1.991 - 4.75, 95% CI; *p*<0.0001); TE: 3.866 (2.254 - 5.56, 95% CI; *p*<0.0001)) (**Fig. 4l**). Taken together these results support the conclusion that chronic pontine myelin injury drives diverse persistent PCS following repeated blast-mTBI.

## Discussion

We showed that brainstem nuclei are vulnerable to a single diffuse blast-mTBI. Within 15 minutes, we observed significant and consistent entry of blood-borne molecules into the brainstem parenchyma and highly variable entry into the cortex. One hour after mTBI, nearly 30% of experimental animals showed neuronal injury, seen as uptake of extravasated blood-borne dyes into axonal projections and cell bodies, whereas <5% of mice showed similar cellular injuries in the cortex. By four hours post-injury, nearly 50% of blast-mTBI mice showed cortical neuronal injuries. This suggests that a therapeutic window to prevent widespread cortical dissemination may exist in the initial 1 to 4 hours following blast-mTBI. Supported mechanisms include subtle neuroinflammatory processes that are likely critical determinants of brain-wide secondary injury, such as swelling of the brain. Such swelling develops within 14 days, is typically seen as significant ventricular enlargement, and is often mistaken for hydrocephalus.^46^ However, when single mTBI-injured rodents are allowed to recover for a month, we found no overt microgliopathy or myelinopathy (**Fig. 3i**); this suggests that repeated injury is the key driver of these phenotypes.

The way in which repeated TBIs provoke the development of chronic pathology and persistent PCS is not well understood. We tested whether repeated diffuse mTBI causes the progressive disruption of brainstem structures responsible for changes in arousal and sensation. Using a data-driven approach, we agnostically identified injured areas by surveying the distribution of spatially-resolved, disease-associated microglial phenotypes across pontine nuclei. As expected, we observed that disease-associated microglia respond to neuronal and myelin injuries following the acute-injury patterns marked by microvascular extravasation and diffuse axon injury. We discovered, however, that release of oligodendroglial nuclear IL33 predicts the regional density of disease-associated microglia, which can appear as single cells and small clusters, or “nodules”, previously reported to facilitate the removal of degenerating axons and myelin.^31,47^ Persisting pathologies of the pontine RF, first identified in our mouse model, were subsequently verified in living veterans with repeated blast-mTBI and in neuropathological studies in veterans with blast-mTBI from four independent laboratories.

We observed several highly stereotypical injury patterns to specific pontine nuclei following repeated blast-mTBI. Among these, a key brainstem area critical for good sleep hygiene is the pontine reticular formation. In living veterans with chronic repetitive blast-mTBI, the myelin integrity of the combined rostral and cerebellar pontine projections that were reconstructed by DTI tractography, were significantly correlated with the severity of persistent PCS including sleep impairment. Projections of the ascending RF strongly innervate central thalamic neurons^48,49^ that modulate the depolarization and firing of neocortical circuits through input to the anterior forebrain mesocircuit and frontoparietal network.^50^ These projections thereby affect the organization of goal-directed behaviors^50^ and modulate arousal levels affecting cognition, stress, and sleep. ^48,51,52^ Likewise, the pontine projections to the cerebellum integrate cerebellar function with diverse brainstem nuclei, including most raphé nuclei, the locus coeruleus, the pedunculopontine nucleus, and several segments of the RF.^53^ Although lesions and DTI abnormalities of the SCP have been frequently reported across a variety of TBI mechanisms of injury and severity of TBI,^17,54,55^ their role in the development of persistent PCS remains unknown. What is known is that experimental lesions of the SCP in cats result in nightly sleep loss,^56^ whereas cerebellectomized cats display increased drowsiness without sleep loss;^57^ this suggests that the prominent cerebellar injuries caused by TBI may also be a significant source of sleep morbidity. Lastly, in keeping with our hypothesis that pontine myelin injury is a causal mediator of persistent PCS, as opposed to the injury of specific output tracts which were not declared *a priori*, we did not evaluate the individual contributions of rostral versus cerebellar projections. Subsequent study is needed to determine the relative contributions of TBI-induced injuries to the SCP versus the pontothalamic projections in the development of persistent PCS, since each may pose unique diagnostic and interventional opportunities. Taken together, the results of this study indicate that diffuse traumatic brain injuries caused by blast overpressures may provoke common, though diverse, neurobehavioral symptoms caused by chronic pontine white matter injuries in nuclei subserving their function.

## Methods

### Study Design

The aim of this study was to determine the frequency and pattern of diffuse axonal and white matter injury in the brainstem following blast-mTBI and to evaluate its relationship with common persistent PCS in both a mouse model of diffuse blast-mTBI with attenuated head movement and in military veterans with history of blast mTBI. Blast exposures occur from all directions, and may not be accompanied by any focal blunt force trauma. This study in a military veteran cohort with primarily repetitive blast-mTBI enabled us to examine the effect of repetitive diffuse blast mTBI on the relationship between brainstem structure and persistent PCS that may be generalizable across mTBI without the confounds associated with excessive head movement, which is well established to injure brainstem centers following TBI caused by diverse forms of trauma.

A total of 39 male veterans reporting from 0 to 102 blast-induced mTBIs were studied. Although female veterans were eligible for study inclusion, none enrolled. Study inclusion criteria included documented military service in Iraq and/or Afghanistan with the US Armed Forces during Operations Enduring Freedom, Iraqi Freedom, and/or New Dawn. Veterans meeting VA/Department of Defense/American Congress of Rehabilitation Medicine criteria for mTBI following at least one blast exposure were included in the TBI group.

Exclusion criteria for the study included a history of moderate to severe TBI, seizure disorder, insulin-dependent diabetes, DSM-IV diagnosis of alcohol or other substance use disorder, dementia, bipolar affective disorder, and psychotic disorders. Subjects using medications likely to affect cognitive or behavioral assessments, such as opiates, benzodiazepines, and sedating antihistamines, were also excluded. MRIs were contraindicated for veterans with retained shrapnel or other metal objects, who were thus excluded from MRI.

A total of 232 male wild-type C57BL/6J, 20 Cx3cr1^EGFP^ /CCR2^RFP^ and 15 Cx3cr1tm2.1^cre/ERT2^ / Rpl22tm1.1^Psam/J^ mice age 3 to 6 months were studied. Mice were randomly assigned to either TBI or sham control groups. Group sizes were based on pathology in this and other established TBI models.^17,58-60^ In each experiment, mice from the control and TBI groups were analyzed under identical conditions.

### Human subjects

Human studies were approved by the VA Puget Sound Health Care System Institutional Review Board. All living veteran participants provided written informed consent prior to any study procedures. The study conformed to institutional regulatory guidelines and principles of human subject protections in the Declaration of Helsinki. Veterans with and without mild TBI were characterized by physical, neurological, and behavioral examinations that included assessments for PTSD using the PTSD Checklist-Military version total score, for sleep disturbance using the PSQI total score, and for current post-concussive symptoms using the Likert-scaled NSI+ symptom questionnaire. The NSI+, based on the standard NSI, is a self-report instrument consisting of the base 22 NSI questions plus the following six TBI-associated PCS: 1) disinhibition, 2) apathy/withdrawal, 3) ringing in the ears, 4) mood swings, 5) getting into fights, and 6) slowness in speech. Responses are scored on scale of 0-4; (0) None, (1) Mild, (2) Moderate, (3) Severe, and (4) Very Severe. The Structured Clinical Interview for DSM-IV was used to establish diagnoses of Axis I psychiatric disorders. A lifetime history of TBI was obtained using a semi-structured TBI interview performed by two expert TBI clinicians simultaneously.^61^ Magnetic resonance DTI was performed within 3 months of clinical evaluation.

### Clinical neuropathology

Human donor research brain specimens and their related clinical/exposure information were collected and used as de-identified materials in accordance with procedures approved, as appropriate, by the VA Puget Sound Health Care System, University of Washington, and the Uniformed Services University Institutional Review Boards. Tissue samples were obtained from brains donated, with informed consent by their legal next of kin, of veterans with a history of mTBI caused by blast exposure and control subjects matched by age and sex but without a lifetime history of TBI. Formalin fixed paraffin embedded (FFPE)-brain sections were de-paraffinized and rehydrated using previously described procedures,^17^ blocked in 10% normal goat serum (1 hour at room temperature), and stained overnight at 4°C using the following antibodies: rabbit polyclonal anti-GFAP (Abcam, Burlingame, CA; 1:1,000), mouse monoclonal anti-Iba-1 (Wako Chemicals, Richmond, VA; 1:1,000), or mouse anti-CD68 (Agilent, Santa Clara, CA; 1:100). Heat-mediated antigen retrieval was performed in citrate buffer (pH 8.0; 80°C) for 30 minutes and then cooled to room temperature before further use. Stained slides were mounted with ProLong Diamond antifade mountant (ThermoFisher Sci, Waltham, MA).

### Diffusion Tensor Imaging (DTI)

Magnetic resonance DTI was performed using an established protocol^17,61^ on a 3.0 T Philips Achieva whole-body scanner (Philips Medical Systems, Best, Netherlands) equipped with a 32-channel radiofrequency head coil. Briefly, images were acquired using a single-shot spin-echo echo-planar sequence with TR=10.76 sec; TE=93.5 msec; flip angle=80 degrees; matrix size=128•128; field-of-view (FOV)=256•256; slice thickness=2mm; 64 gradient directions; and b-factors=0 and 3,000s/mm2.

Images were preprocessed to correct for head motion, eddy current, and B0-field inhomogeneity-induced geometric distortion using the Oxford FMRI Software Library (FSL) DTI toolbox. Image slices with large within-slice intensity differences, wrapping abnormalities, or other artifacts were identified by analysis with DTIPrep and subsequently removed. DTI tractography was used to test for correlations between DTI parameters, the number of reported TBIs, and symptom areas related to known brainstem functions. To conduct tractography, DICOM-formatted diffusion images were converted to nrrd data file/header (nhdr) format (http://teem.sourceforge.net/nrrd/format.html) in g-Fortran. Fiber tracts were created in SLICER 4.3.1 (http://www.slicer.org/). After the diffusion data were loaded into Slicer in nhdr format, tensor estimates were created by converting the diffusion weights.

The resulting process created scalar measurements that were used to create corresponding images of FA. The Editor module in Slicer was used to create a label map and a region of interest (ROI) located in the four ROIs in the pons (**Fig. 4d**). Fiber tracts for each individual participant were reconstructed from these seed regions using 1mm spacing thresholds, and the resulting fiber tract vtk file was read into g-Fortran for quantitation of FA, mean diffusivity, radial diffusivity, and axial diffusivity. These values were analyzed using nonparametric Spearman correlation statistical analysis with respect to the log10(number of reported TBIs + 1), hours of self-reported nightly sleep, and other symptoms (**Fig. 4**), with final reported p-values controlled for multiple comparisons using a rigorous 1% false discovery rate (FDR) adjustment. Each volume of interest (VOI) had a volume of approximately 20mm^3^ and was manually positioned. Evaluation of pontine DTI averaged bilateral VOIs for rostral pontine (RP) and pontocerebellar (PC) tracts with Montreal Neurologic Institute atlas x, y, z coordinates of (Left RP [-27.1, -20.0, -6.0]), (Right RP [5.0, -20.0, -6.0]), (Left PC [-30.0, -38.0, -33.0]), and (Right PC [5.0, -38.0, -33.0]; exemplars are found as white boxes in **Fig. 4d**).

### Animals

Mice used in this study comprised group-housed, 3-to-6-month-old male C57Bl/6J wildtype, Cx3cr1^EGFP^ /CCR2^RFP^ [B6.129(Cg)-Cx3cr1tm1Litt-Ccr2tm2.1Ifc/JernJ (JAX stock # 032127)], and microglial RiboTag mice [heterozygous offspring of B6.129P2(C)-Cx3cr1tm2.1(cre/ERT2) mice (JAX stock # 020940)] crossed with RiboTag flox mice (JAX stock # 011029 B6N.129-Rpl22tm1.1Psam/J)). Animals were maintained on a 12-hour light/dark cycle with *ad libitum* food and water access. All study procedures were in accordance with Association for Assessment and Accreditation of Laboratory Animal Care guidelines and approved by the VA Puget Sound Health Care System Institutional Animal Care and Use Committee.

### Modeling blast overpressure

TBI was modeled with a shock tube designed to simulate open-field blast explosions (Baker Engineering and Risk Consultants, San Antonio, TX) as described elsewhere.^12,17^ Briefly, anesthetized mice were maintained on a non-rebreathing anesthesia apparatus (2% isoflurane in 1 Lpm oxygen) while secured on a gurney with their ventral side oriented toward the oncoming blast overpressure wave and then placed into the shock tube. TBI mice were paired with sham control mice and both were similarly secured in the shock tube while under anesthesia for an identical amount of time. Mice received blast or sham procedures once per day, with 3x treatment mice receiving one treatment per day for three days. The mean overpressure wave characteristics for 102 blast overpressure waveforms generated in this experiment are 20.62+/-0.15psi; 5.65+/-0.036ms; 0.038+/-0.00019psi·ms, including peak intensity (psi), initial pulse duration (ms), and impulse (psi·ms), respectively. **Figure S2** displays an average of waveforms from the study; this image, taken at random, consists of 102 overpressures and is consistent with the accepted properties of mild to moderate blast exposure,^17,62,63^ resulting in a >95% survival rate. After TBI and sham exposures, mice were monitored and generally regained normal appearance within 4 hours of exposure.

### Tamoxifen treatments

Three weeks after TBI or sham treatments, gene recombination was induced in microglial RiboTag mice by two i.p. injections of tamoxifen (4mg in 200µl corn oil [Sigma, St. Louis, MO; C8267]) that were administered two days apart.

### RiboTag immunoprecipitation

Tamoxifen-treated Cx3cr1creER-Rpl22 RiboTag mice were deeply anesthetized with a lethal injection of sodium pentobarbital (210 mg/kg, i.p.) followed by transcardial perfusion with PBS-containing cycloheximide (100µg/ml). Brains were removed; subdissected on ice into brainstem, cerebellum, and cortical hemisections; weighed; flash frozen; and stored at -80°C until used. Upon use, approximately 20 to 30mg of each frozen tissue was cut and collected into RNase-free tubes followed by homogenization by hand with a tube-fitting disposable pestle using 2-3% weight per volume of the following buffer (HB-S): DTT (Sigma, St. Louis, MO; 646563, 1mM), Protease Inhibitor Cocktail (Sigma, St. Louis, MO; P8340, 1x), RNAsin (Promega, Madison, WI; N261B, 200 units/ml), cyclohexamide (Sigma, St. Louis, MO; C7698, 100µg/ml), heparin (Sigma, St. Louis, MO; H3393, 100 mg/ml) in RNase-free deionized water. Homogenized samples were centrifuge-clarified (12,000 x g, 10min, 4°C), and supernatants were transferred into fresh RNase-free tubes followed by incubation (4°C for 4 hours with tube rotation) with mouse monoclonal anti-HA.11 (Covance, MMS-101R, 3 uL/sample). After 4 hours, 200uL of Protein A/G Magnetic Beads (Promega, Madison, WI; 88803) were added to each sample and rotated at 4°C overnight. Beads were washed and pre-equilibrated in homogenization buffer and blocked with 4% bovine serum albumin (Sigma, St. Louis, MO; 3117332001) for 1 hour prior to use. The next day, mRNA transcripts and their associated antibody-bound ribosomes were precipitated by magnet and washed (3 × 15 minutes each, 4°C with rotation) in 1 ml high salt buffer (500 mM Tris pH 7.4, 300 mM KCl, 12 mM MgCl_2_ 1% NP-40, 1 mM DTT, 100 µg/ml cyclohexamide in RNase-free water). Immediately after the final wash, transcripts were dissociated from their precipitating complexes by addition of RLT Buffer (Qiagen, Germantown, MD; 79216) supplemented with 10 µl/ml RNase-free β-mercaptoethanol (Sigma, St. Louis, MO; 63689). RNA was purified by RNeasy Plus Micro kit (Qiagen, Germantown, MD; 74034) according to manufacturer’s protocol, followed by RNA quantification using a Nanodrop spectrophotometer (Thermofisher Sci, Waltham, MA). cDNA of precipitated transcripts was made using the SuperScript VILO cDNA synthesis kit (Thermofisher, Waltham, MA; 11754050) according to the manufacturer’s protocol. Yield of cDNA preparations was determined by nanodrop (input: ∼5ng/µl; immunoprecipitant: ∼1.5-2.5 ng/µl). To verify enrichment of mouse microglial genes, immunoprecipitants were analyzed in triplicate by real-time PCR on a StepOnePlus Real-Time PCR System (Thermofisher, Waltham, MA), using KAPA Universal SYBR Green master mix and the following primers: Aif1(NM019467.3) forward 5’-GGATTTGCAGGGAGGAAAAG, reverse 5’-TGGGATCATCGAGGAATTG; NTRK2 (NM008745.3) forward 5’-TGTTGCCTATCCCAGGAAGTG, reverse 5’-CTGCAGACATCCTCGGAGATTA; GFAP (NM010277) forward 5’-ACCATTCCTGTACAGACTTTCTCC, reverse 5’-AGTCTTTACCACGATGTTCCTCTT; GAPDH (NM008084) forward 5’-CTGCACCACCAACTGCTTAG, reverse 5’-ACAGTCTTCTGGGTGGCA GT. Representative results are shown in **Figure S3**. The following thermocycler settings were used: 95°C for 20 seconds followed by 40 cycles of denaturation (95°C for 3 seconds), primer annealing and extension (60°C, 30 seconds) followed by a melting curve. Gene expression was normalized within subjects to GAPDH levels and quantified using the 2^-ΔΔCT^ method. ROX was used as a reference dye.

### mRNA preparation and analysis

RiboTag-immunoprecipitated mRNA was ribosomal RNA depleted using the Ribo Zero Gold Magnetic system (Epicenter/Illumina, San Diego, CA). We prepared RNA-sequencing libraries using ScriptSeq v2 kit (Epicenter) followed by single-end sequencing (1x50bp) on an Illumina HiSeq 2500, generating 4 × 10^6^ mean reads per sample. RNA-seq fastq files were aligned to the mouse genome build *mm10* with *STAR* aligner and processed into transcript-level summaries using the expectation maximization algorithm *RSEM*. Transcript-level summaries were combined into gene-level summaries by adding all transcript counts from the same gene. Gene counts were normalized across samples using *RLE* normalization, and the gene list was filtered based on mean log_2_ (counts per million reads) > 4 in at least 3 samples in any group, which left about 14,000 “detected” genes for further analysis. Consistency of replicates was inspected by principal component analysis using *R*. Differential expression was assessed through sequential analyses of both an initial discovery cohort to identify candidate genes (α=0.05) and a secondary validation cohort (α=0.05). The candidate differential genes, defined as the shared set of overlapping genes, was further evaluated. Forty-percent of the identified candidate differentially expressed genes have been previously reported (**Table S1**).

Gene set enrichment analysis (GSEA) was conducted in DAVID (https://david.ncifcrf.gov/tools.jsp) using the full list of candidate differentially expressed genes for brainstem. Official gene symbols were queried against the *Mus musculus* species using the default settings for analyses conducted by DAVID. Gene Ontology terms with Benjamini-corrected *p*<0.05 were reported.

## Data availability

Data are available through reasonable requests addressed to the corresponding author.

### Neurovascular permeability studies

#### Radiolabeled tracer preparation

Albumin (Sigma, St. Louis MO) was radiolabeled using established protocols^64^ with ^99m^ Tc (GE Healthcare, Piscataway, NJ). Hydrochloric acid was used to pH-adjust a solution of aqueous stannous tartrate (240 mg/ml) and albumin (1 mg/ml) to a final pH of 3.0 before the addition of one millicurie of ^99m^ Tc-NaOH4 to the mixture, which was then allowed to react for 20 min. The resulting radiolabeled ^99m^ Tc-albumin was column-purified (G-10 Sephadex; GE Healthcare, Piscataway, NJ) in 0.1 ml fractions of phosphate buffer (250 mM). Purified ^99m^ Tc-albumin was > 90% acid precipitable in 1% bovine serum albumin and trichloroacetic acid (30%) (1:1 mixture). Each mouse was administered 5 × 10^6^ counts per minute (cpm) of purified ^99m^ Tc-albumin fraction in 0.2 ml lactated Ringer’s solution containing 1% BSA. ^14^ C-sucrose was supplied as an ethanol-dissolved product (GE Healthcare, Piscataway, NJ). Ethanol was evaporated before use and resuspended at 1 × 10^7^ cpm in 1% BSA lactated Ringer’s solution. <u>Radiolabeled tracer injections:</u> At 15 minutes, 24 hours, or 72 hours after mTBI or sham treatments, mice were anesthetized with urethane (4 g/kg; 0.2 ml; i.p.) followed by exposure of the jugular vein and injection with ^14^ C-sucrose or ^99m^ Tc-albumin in 0.2 ml of lactated Ringer’s solution with 1% BSA for 10 minutes. The descending abdominal aorta was cut to collect blood. Vascular contents of the brain were cleared by left cardioventricular perfusion (20 ml lactated Ringer’s solution per minute) after clamping the descending thoracic aorta and severing both jugular veins. Two brains with incomplete blood washout were excluded from analysis. After perfusion, the brain was removed; subdissected into cortical hemispheres, brainstem, and cerebellum; and individually weighed. The analytical results of these experiments with respect to cerebellum were recently published by our group.^64^ Radioactivity was calculated as cpm/g tissue or cpm/ml serum, as appropriate. The brain tissue radioactivity was then divided by the corresponding serum radioactivity to yield units of microliters/gram of brain tissue.

### *In vivo* dextran labeling of injured cells

Extravasation of blood-borne dextran to label coincident vascular and neural injury was accomplished as described elsewhere.^17^ Briefly, following induction of isoflurane anesthesia prior to TBI or sham procedures, mice were injected with 100 μl of 400 mg/ml 10kDa dextran labeled with tetramethylrhodamine (Life Technology, Grand Island, NY) into the retro-orbital sinus. After TBI or sham treatments, mice then recovered for 1 or 4 hours before euthanasia and transcardial perfusion with phosphate-buffered saline followed by 4% formalin.

### Animal MRI

MR imaging was acquired on 12 mice (6 sham and 6 single mTBI) at 24 hours post-injury and again at 30 days post-injury. Mice were anesthetized with isoflurane and scanned over the entire brain using a high-resolution 14 T MRI (Avance III, vertical bore, Bruker BioSpin Corp, Billerica, MA). T2 quantitative mapping (T2) was acquired with a voxel size of 0.12 × 0.12 × 1.0 mm^3^, 15 slices, TR 1⁄4 2000 ms,16 echoes, spacing: 6.7 ms, TE Effective 1: 6.7 ms, TE Effective 2: 13.4 ms, and was used to measure ventricular volume and scan for evidence of parenchymal bleeding within visible areas. All analyses were performed by a blinded investigator (DJC).

Analysis of T2 quantitative maps required threshold bounding set from 36 to 200, which included ROIs but excluded normal cortex and white matter. Manually drawn ROI analysis was performed using Image J software. ROIs included the volume of voxels within the threshold boundary comprising the lateral, 3^rd^, and 4^th^ ventricles, across a total of seven image slices, and total volumes were calculated by multiplying with voxel size in mm^3^.

### Mouse sleep assessments

Noninvasive measurements of sleep bout duration and the sleep:wake ratio were acquired using an automated sleep/wake scoring system designed for rodents (PiezoSleep; Signal Solutions, Lexington, KY). One week after the final TBI or sham procedure and again three months later, mice were placed in open-floored chambers installed with piezoelectric sensors are designed to detect and track the previously characterized sleep activity of mice.^65^ Mice were allowed to acclimate for 2 days before their undisturbed activity was recorded over the next 3 days following previously established methods.^37,66^ Data were analyzed using the Sleep Statistics Toolbox (Signal Solutions, Lexington, KY).

### Microscopy

Microscopy was performed as described elsewhere.^17^ Briefly, brains were post-fixed in 10% neutral buffered formalin at 4°C for 3 to 5 days followed by 24 hours of equilibration in 30% sucrose/PBS prior to embedding in OCT (Tissue-Tek, Torrance, CA). Antigen retrieval was performed using 50 mM of sodium citrate (pH 8.0) with heat (80°C, 30 min). Cryopreserved floating tissue sections were permeabilized with 1% Tx-100 (Sigma, St. Louis, MO), blocked with 10% bovine serum albumin (1 hour, room temperature), and immunostained and mounted using Prolong Gold Antifade Reagent (Thermofisher, Waltham, MA). The following antibodies were used overnight at 4°C: mouse anti-CNPase clone SMI-91 (Biolegend, Dedham, MA; 1:1,000), astrocyte marker anti-GFAP (Millipore, Billerica, ME; 1:1000), neuronal marker anti-neurofilament-heavy chain (Nf-HC) (Aves, Tigard, OR; 1:1000), mouse monoclonal TauC3 (ThermoFisher, Waltham, MA; 1:1000), rabbit anti-phospho Tau 396 (AnaSpec, Freemont, CA; 1:1000), and mouse monoclonal anti-Iba-1 (Wako Chemicals, Richmond, VA; 1:1,000). Corresponding secondary antibodies were applied for 2 hours at room temperature (Jackson Immunoresearch, West Grove, PA; 1:1,000). Confocal microscopy was performed with a Leica TCS SP5 II with tunable emission gating. Representative brainstem regions imaged for analyses are identified in **Supplemental Figure 5**. To quantify the degree of myelin internalization by microglia, z-plane images of tissue areas (approximately 600 µm x 600 µm x 50 µm) were acquired by performing sequential, between-stack, single-photon excitation at 488 nm and 543 nm using system optimized stepping (∼2 µm). Images were acquired with the Leica Application Suite and processed using linear contrast and brightness adjustments applied identically and simultaneously to all data being directly compared within each experiment.

### Imaging mass cytometry

Metal-conjugated antibodies were either purchased (Standard Biotools, San Francisco, CA) or custom-made by conjugation using the MaxPar X8 labelling kit (Standard Biotools, San Francisco, CA) and carrier and preservative-free antibody formulations following the manufacturer protocol. Typical antibody recovery was 59 ± 0.16%. Tissue staining with metal-labeled antibodies was similarly accomplished using the manufacturer’s protocol for formalin-fixed paraffin-embedded slides with minor modifications. Briefly, tissue sections of either formalin-fixed human paraffin embedded tissues or covalently bound, agarose-block embedded mouse brain sections were used for imaging mass cytometry. Paraffin embedding and sectioning followed standard histological procedures, whereas preparation of agarose-block embedded brains followed procedures developed and published by the Allen Institute for Brain Science for use in serial two-photon tissue tomography (dx.doi.org/10.17504/protocols.io.bf65jrg6). One each TBI and sham mouse brain were randomly paired and embedded together into the agarose block. The brains were oriented in parallel such that the sham brain cortex was always oriented facing outward. Agarose block–embedded sections were cut at 50 µm thickness (immediately after aqueous two-photon imaging). Slices cut from agarose block–embedded brains were robotically mounted with a TissueVision 1600FC SlicerPlacer module (Tissuecyte, Newton, MA) onto superfrost plus slides (Thermo Fisher Scientific Bothell, WA) and subsequently air-dried for up to one year before use. DNA intercalator (Iridium 191/193, 201192B; Standard Biotools, San Francisco, CA) was diluted 1:4000 in Dulbecco’s phosphate buffered saline (21600-051, in 18.2 MΩ water; Thermo Fisher Scientific; Bothell, WA) for 30 minutes at room temperature before slides were washed in 18.2 MΩ water, air-dried overnight, and stored at room temperature until used.

Coronal brainstem hemisections were routinely ablated using the Hyperion UV laser system with 1 µm^2^ diameter (Standard Biotools, San Francisco, CA). The ablated materials are carried by helium into the Helios mass cytometer, which uses a time-of-flight mass detector to detect and timestamp the abundance of a user-defined mass range for each rasterized 2-dimensional location.^67^ The Hyperion instrument was tuned daily according to the manufacturer’s protocol.

### Single cell gating, analysis, and image visualization

Imaging mass cytometry datasets were collected in MCD file format and exported as single channel TIFs for bulk import into CellProfiler. There, segmentation of single cells based on the watershed gradient algorithm using DNA intercalator channels (191Ir, 193Ir) and a proprietary mix of plasma membrane markers (channels 195Pt, 196Pt, 198Pt; IMC Cell Segmentation Kit, TIS-00001; Standard Biotools, San Francisco, CA) was performed to define cell nuclei and single cell boundaries, respectively. Microglia were gated by selecting cells with marker intensities for CCR2^-^ (<2.5) and Iba1^+^ (> 6) to delineate them from peripheral immune cells expressing higher levels of CCR2. DAMs were gated by selecting the following parameters: Iba1 > 6, CD68 > 10, CCR2 < 2.5, ApoE > 8, and CD63 >2. The thresholds for CCR2, ApoE, and CD63 positive signals was determined by comparison of the CD63+ microglial cells that were identified by manual observation, relative to other cell types. The threshold for CD68 high or low signal was determined by the comparison to manually identified microglia in sham control animals establishing the lower limit. All major cell types, including astrocytes, neurons, oligodendrocytes, peripheral immune cells, and vascular endothelium, were validated by manual annotation.

Brainstem images were aligned to the Allen mouse brain reference map^68^ and segmented by anatomical region in HistoCAT.^69^ Using HistoCAT, Barnes-Hut t-SNE, and phenograph, neighborhood analyses (using a proximity of 6) were conducted using default settings. Images representing single and merged channels, such as **Fig. 2f**, were saved as TIF files via MCD viewer software (Fluidigm), imported into Adobe Photoshop 2021 (Adobe Systems, San Jose, CA), and processed with a Gaussian blur filter to de-speckle images. Afterward, all images being directly compared within a figure panel were uniformly and simultaneously adjusted to optimal brightness and contrast levels using a single adjustment mask.

### Western blot analyses

Brains were dissected in PBS on ice. Protein lysates were prepared as described elsewhere.^17^ Briefly, phosphatase inhibitor cocktail sets 2 and 3 (Sigma, St. Louis, MO) were added (10 μl/ml) to lysis buffer, and tissues were homogenized twice by hand using an Eppendorf tube-fitting pestle (Eppendorf, Hauppauge, NY) before centrifuge clarification at 12,000xg for 10 minutes. TGX Criterion 4-20% SDS-polyacrylamide gels (Bio-Rad, Hercules, CA) were loaded with 20 μg protein per lane using supernatants of the clarified lysates from individual animals. Membranes were then blocked with 10% nonfat milk (1 hour at room temperature) and probed with antibodies recognizing the following targets: anti-pathological tau clones AT8 and AT270 (Thermo Fisher Scientific, Rockford, IL); anti-mouse tau (total) clone Tau5 and anti-phospho Tau S396 (Life Technologies, Grand Island, NY); MOBP (Genetex, Irvine, CA); CNPase (Biolegend, San Diego, CA); mouse anti-MBP and rabbit antibodies anti-LINGO1, anti-GFAP, and anti-MnSOD (EMD-Millipore, Billerica, MA); and Pyruvate kinase (Rockland, Gilbertsville, PA). Probes for the load control protein, pyruvate kinase, were conducted after stripping.

### Causal mediation analysis

Mediation analysis evaluates the statistical probability of a known third (declared) variable to mediate the primary relationship between independent and dependent variables. The effect of changes in pontine FA (adjusted for age and PCL-M total scores for PTSD symptom severity) to mediate behavioral and physiological symptoms (dependent variables) following self-reported TBI (independent variable) was first evaluated with multivariable linear regression in R (**Fig. 4j**). A subsequent validation analysis was conducted with the R statistical package Mediate using nonparametric bootstrapping procedures and also used age- and PTSD symptom severity-adjusted pontine FA (**Fig. 4k**). This indirect effect is measured as the portion of the relationship between two variables mediated by the third variable.^70,71^ In validation analyses, unstandardized indirect effects were computed thrice for each of 10,000 bootstrapped samples, and the 95% confidence interval was computed by determining the indirect effects at the 2.5th and 97.5th percentiles. Sensitivity analyses were conducted in R and are illustrated in **Figure S5**.

### Statistical analyses

Data are presented as distributions (violin plots) or mean ± standard error of the mean (bar graphs). <u>Multivariable linear regression:</u> Standard analysis of variance (ANOVA) was used for planned statistical tests involving multiple measures or groups. False discovery rate-adjusted post-hoc analyses with alpha = 0.05 were conducted as indicated. T-tests were used for planned comparisons involving two normally distributed groups, with nonparametric Mann-Whitney tests used for unpaired comparison of non-normal distributed groups. Bivariate linear or simple linear regression was used to predict DAM frequency based on IL-33 expression. Multivariate linear regression analysis reported correlations adjusted for age, depression, PTSD symptom severity, sleep apnea, and alcohol use. Statistical significance was defined as *p*≤0.05. Sample sizes were based on previous reports.^17,60,72^ All analyses were determined using Prism 8.4 (GraphPad, San Diego, CA), SPSS software (IBM, Armonk, NY), or R version 3.6.1 (R Core Team, Vienna, Austria).

## Data Availability

After peer-review and subsequent acceptance for publication in a peer-reviewed journal, all data reported in the present study are available upon reasonable request to the authors.

## Author contributions

JSM: Conceptualization, formal analysis, supervision, funding acquisition, investigation, visualization, writing—all drafts, writing—review and editing; AGS, MMC, AFL, CM, AC, JRP, AS-D, and RGT, KFP, DAM, CDK: Formal analysis, investigation; MY: Validation; TLR and KDM: Formal analysis, performed the DTI analyses; DJC and DGC, DRM, JWR, MO: Investigation; DPP, WAB, JPM, MAR, EAC, CLS, CDK, DAM and ERP: Conceptualization, resources.

We’d like to thank Dr. Erica Melief and Aimee Schantz for outstanding administrative support, and Lisa Keene, Amanda Keen, and Katelyn Kern for expert technical assistance, and the incredible generosity of the military Veteran and civilian participants, and their families, without whom this work would be impossible.

---

**Table S1.**
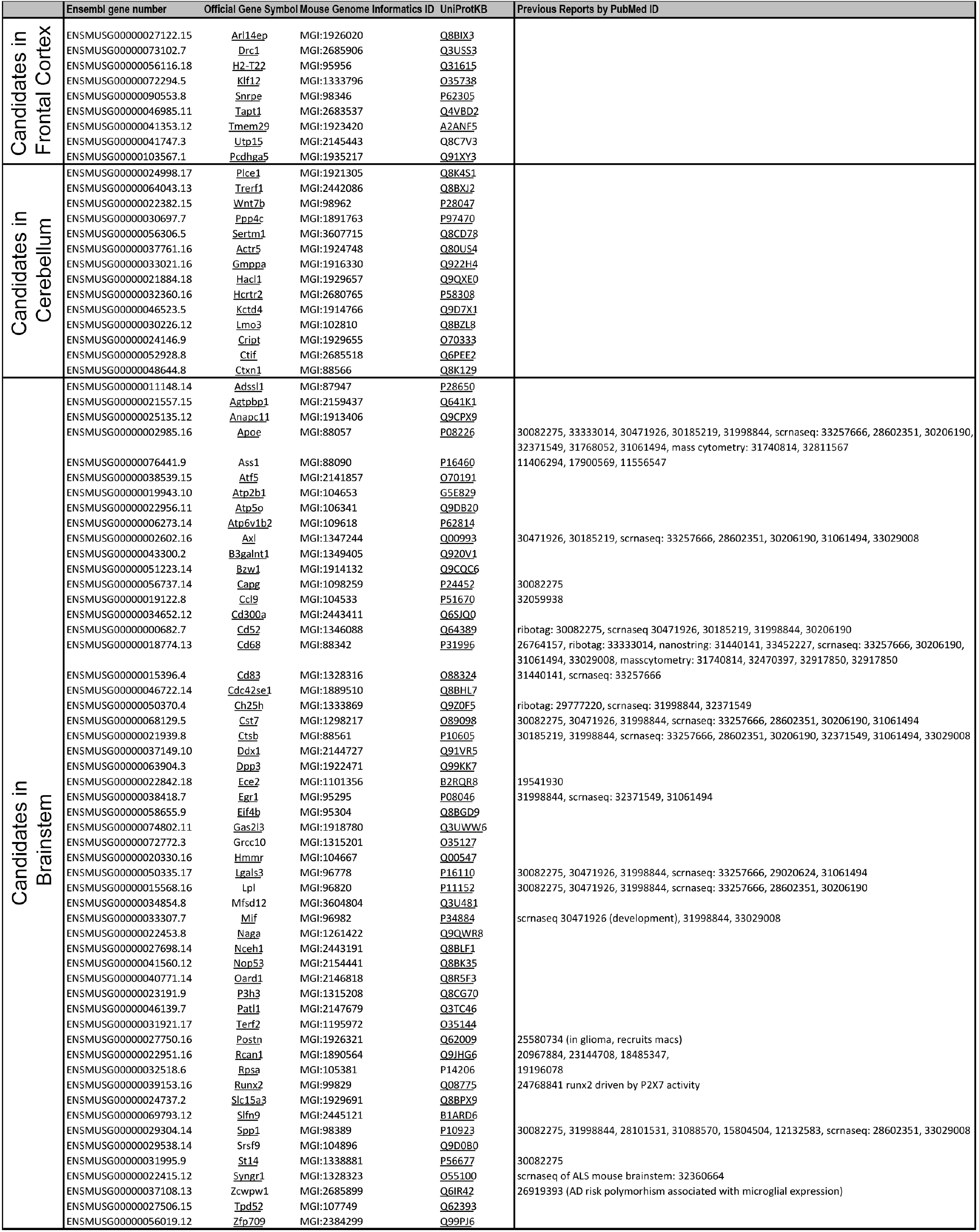
Table of candidate DEGs previously reported.

**Table S2.**
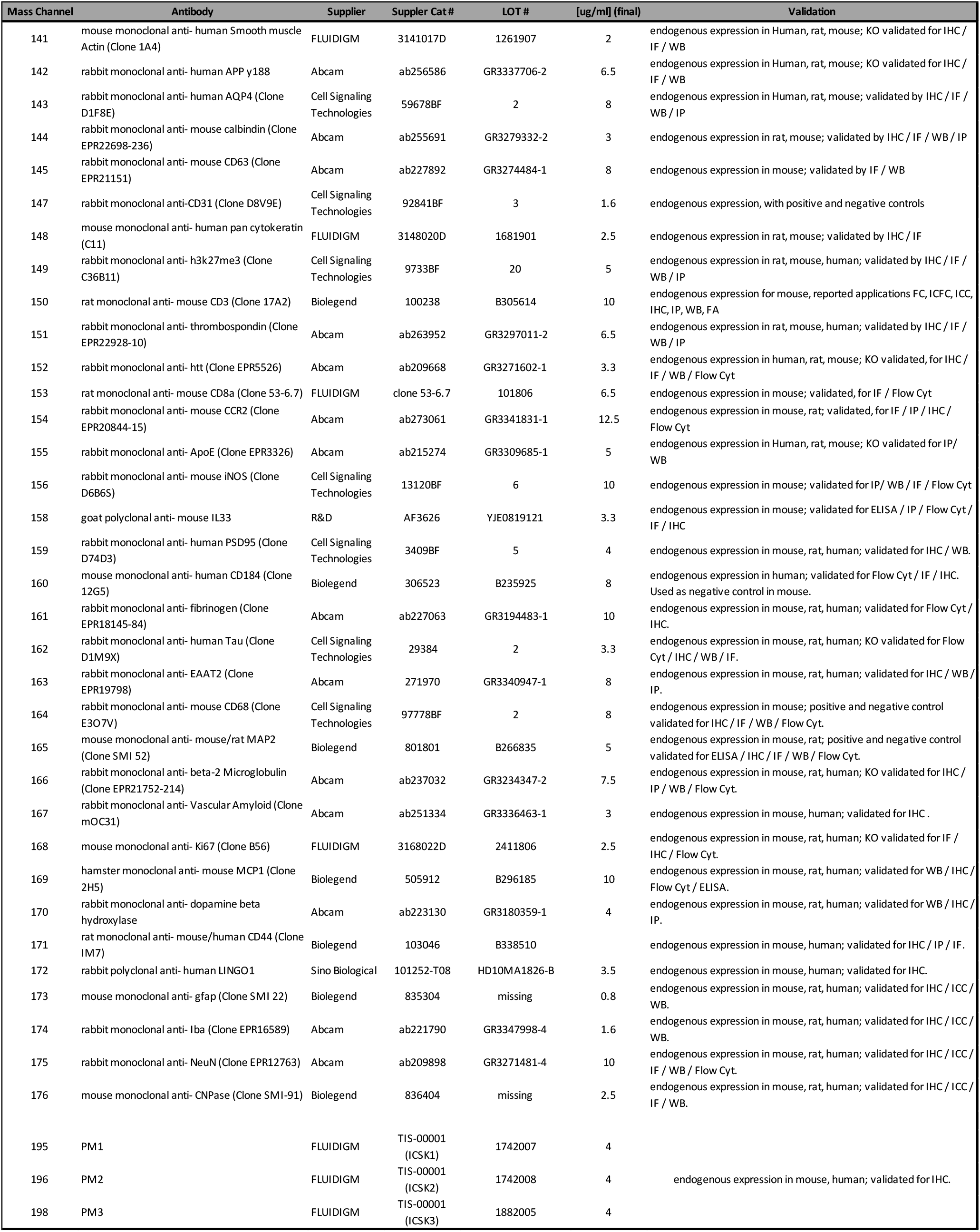
Table of imaging mass cytometry antibodies.

**Supplemental Table 3.**
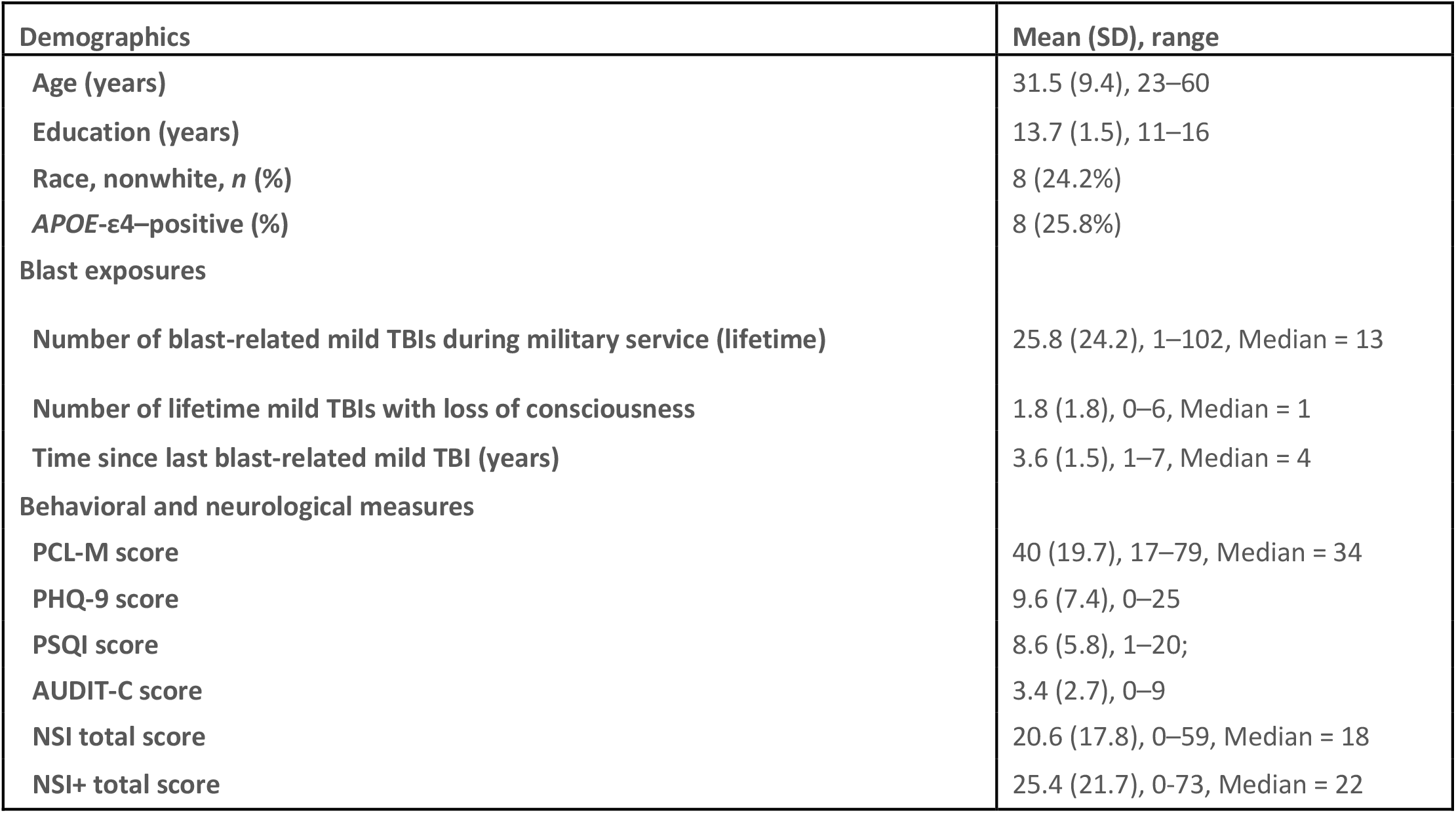
Subject Demographics

**Figure S1.**
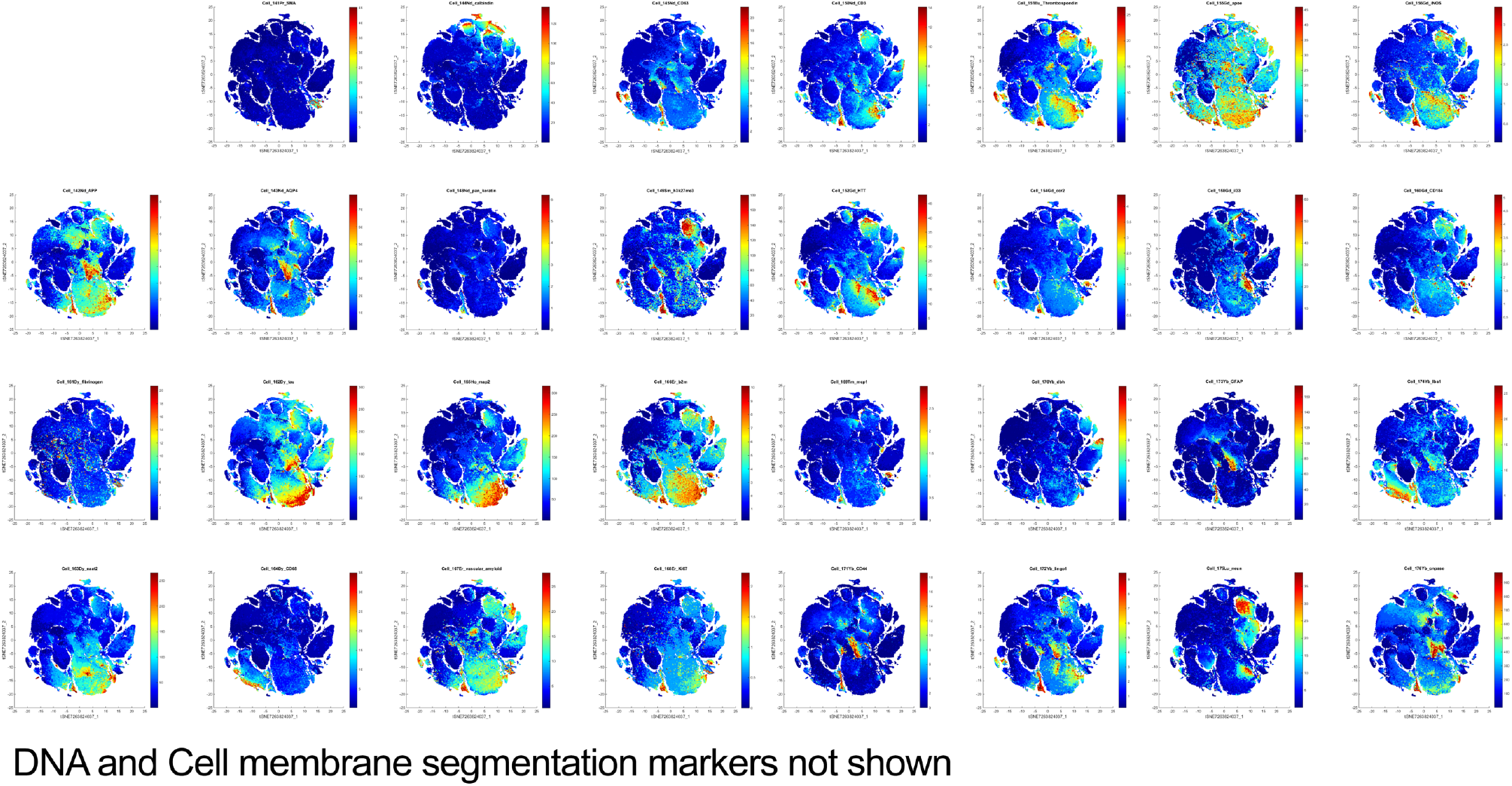
T-SNE plots for separate IMC mass channels included in the study.

**Figure S2.**
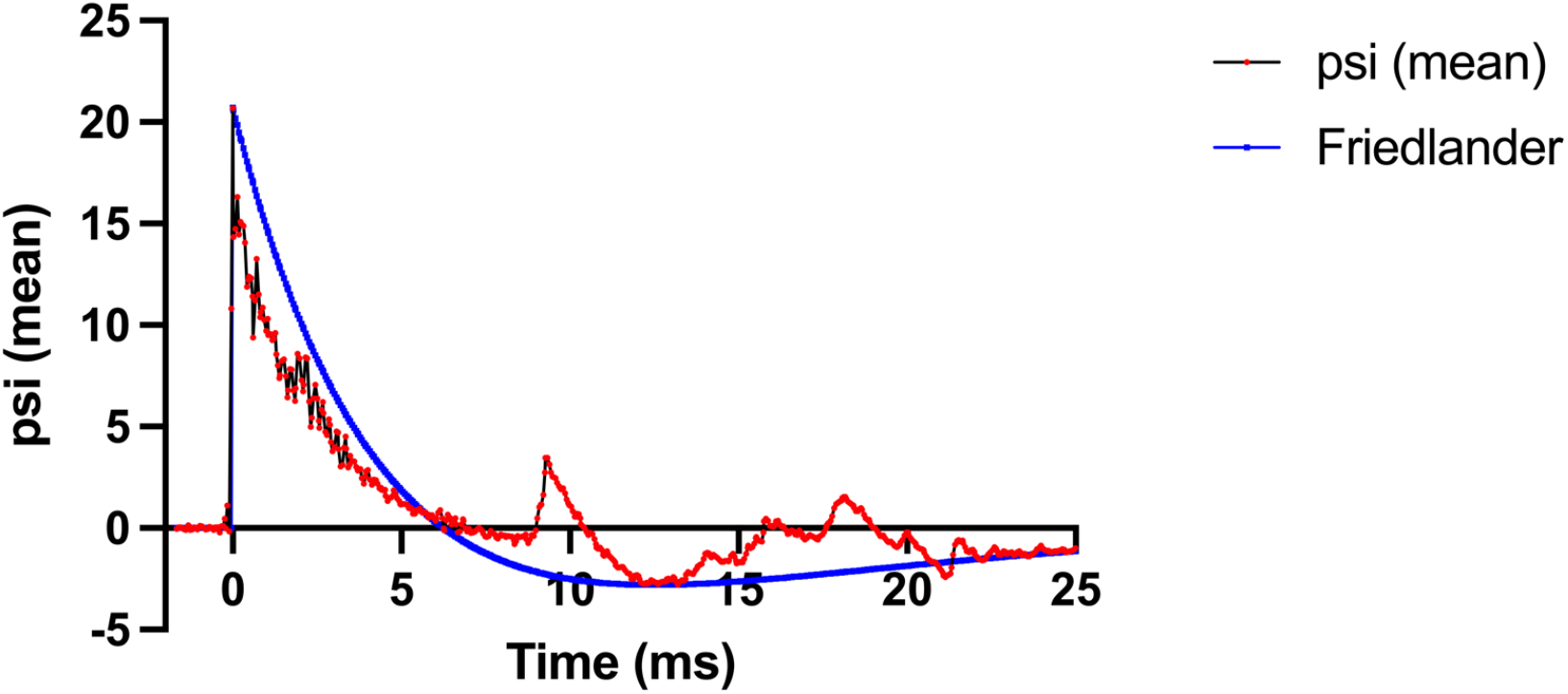
Average overpressure waveform for diffuse mTBI model. Data represent the average timestamped pressure measurements taken 5 cm above the animal over 102 trials occurring over several days (red shown with black bars indicating ±SEM). The blue line represents the idealized Friedlander waveform produced by 11.35 kg of trinitrotoluene (TNT) at a distance of 6.2 m.

**Figure S3.**
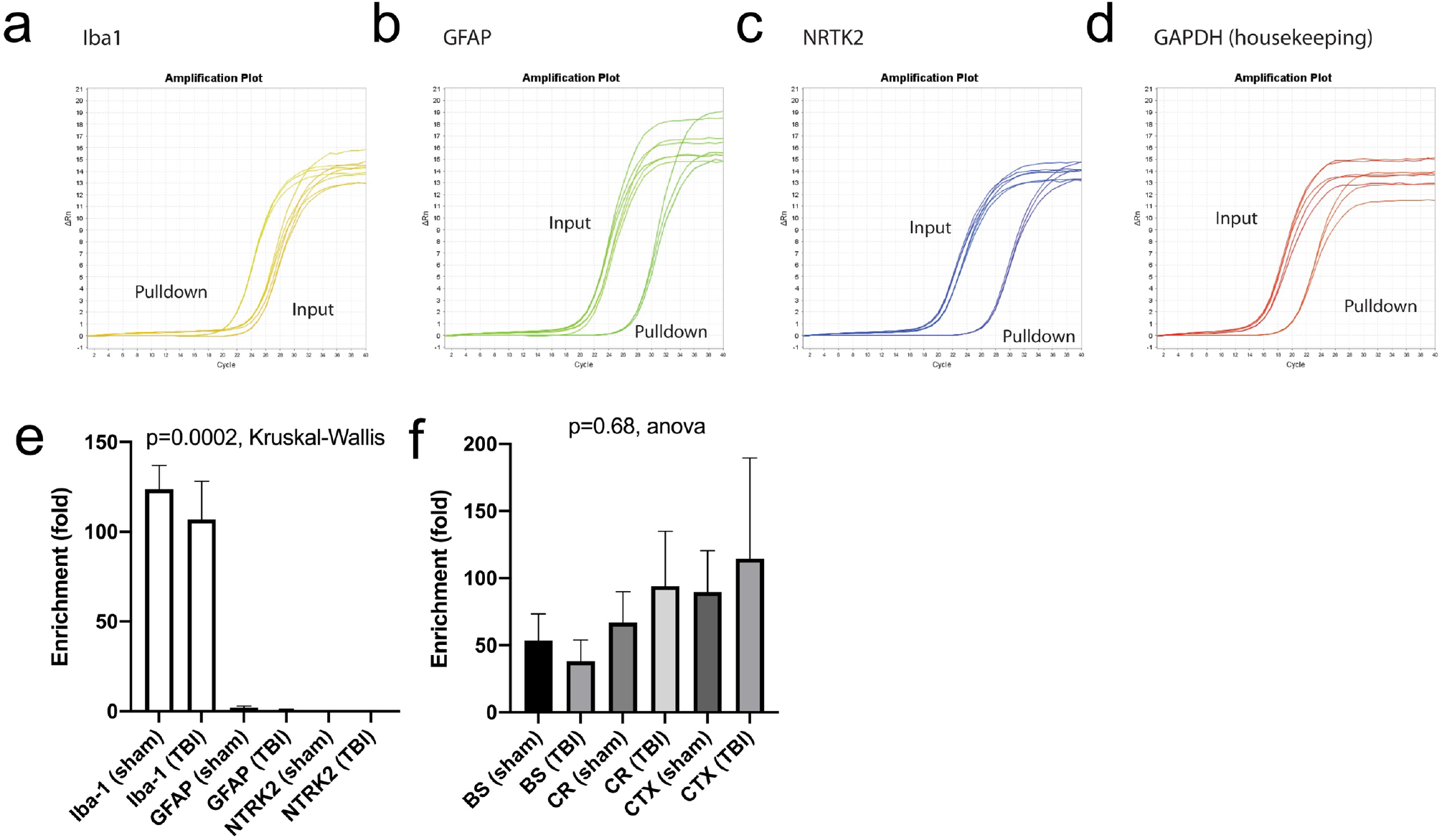
Analysis of mRNA enrichment by RiboTag immunoprecipitation. **(a-d)** Real-time (RT) PCR amplification curves of **(a)** microglial Iba-1, **(b)** astrocytic GFAP, **(c)** neuronal NTRK2, and **(d)** the ubiquitous housekeeping gene GAPDH. Each curve represents either immunoprecipitated mRNA or input samples from individual mice. **(e)** Quantification of RT-PCR enrichment analyses using cortical hemisection tissue (N=4-8 per tissue). **(f)** Relative Iba-1 enrichment across brainstem (BS), cerebellum (CR), and cortical tissues (CTX) (N=2-6 per tissue).

**Figure S4.**
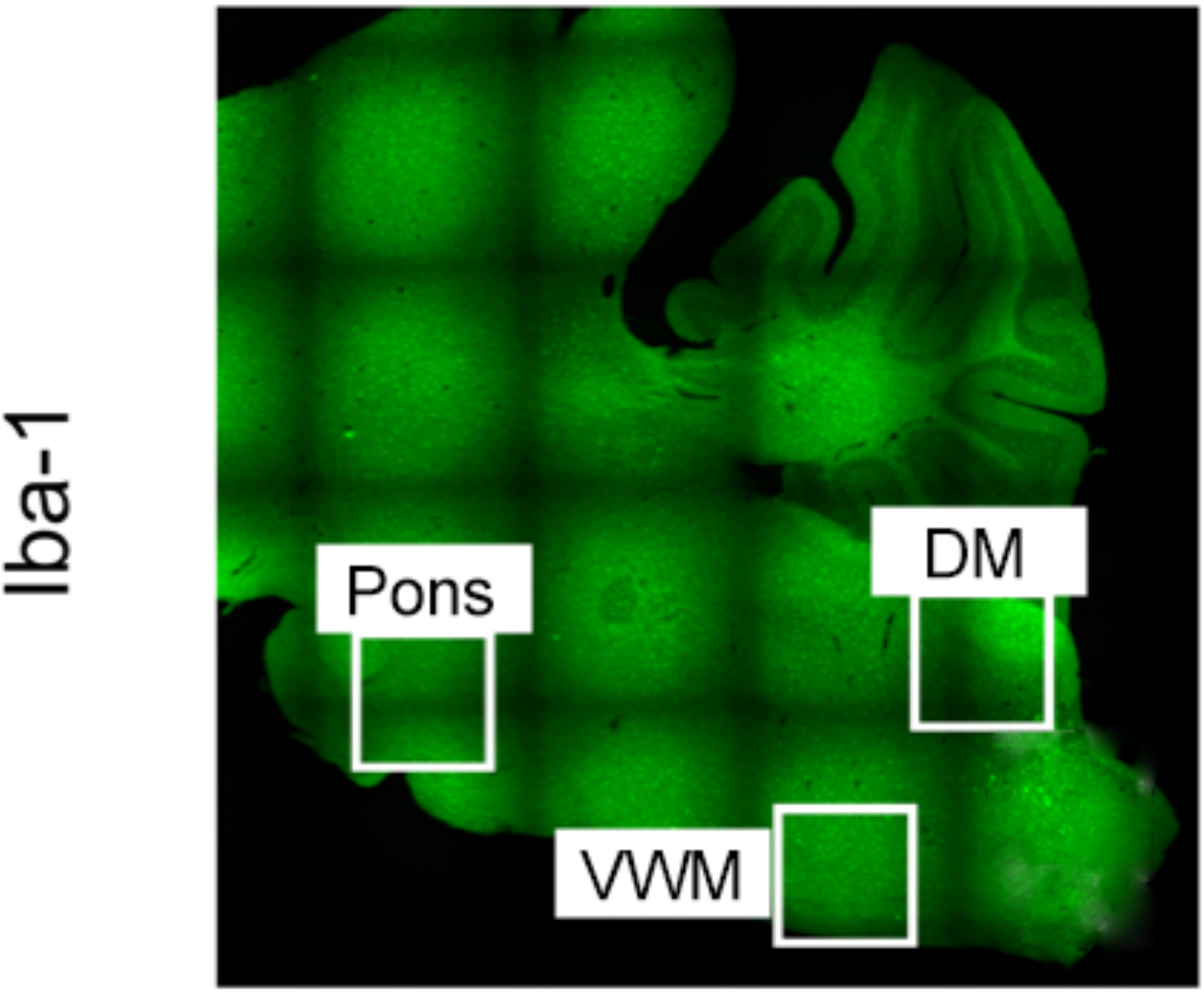
Locations used for confocal microscopy analyses of microglia internalization of myelin CNPase. Sagittal mouse brain section shows location of images taken from brainstem pons, dorsal medulla (DM), and ventral white matter (VWM).

**Figure S5.**
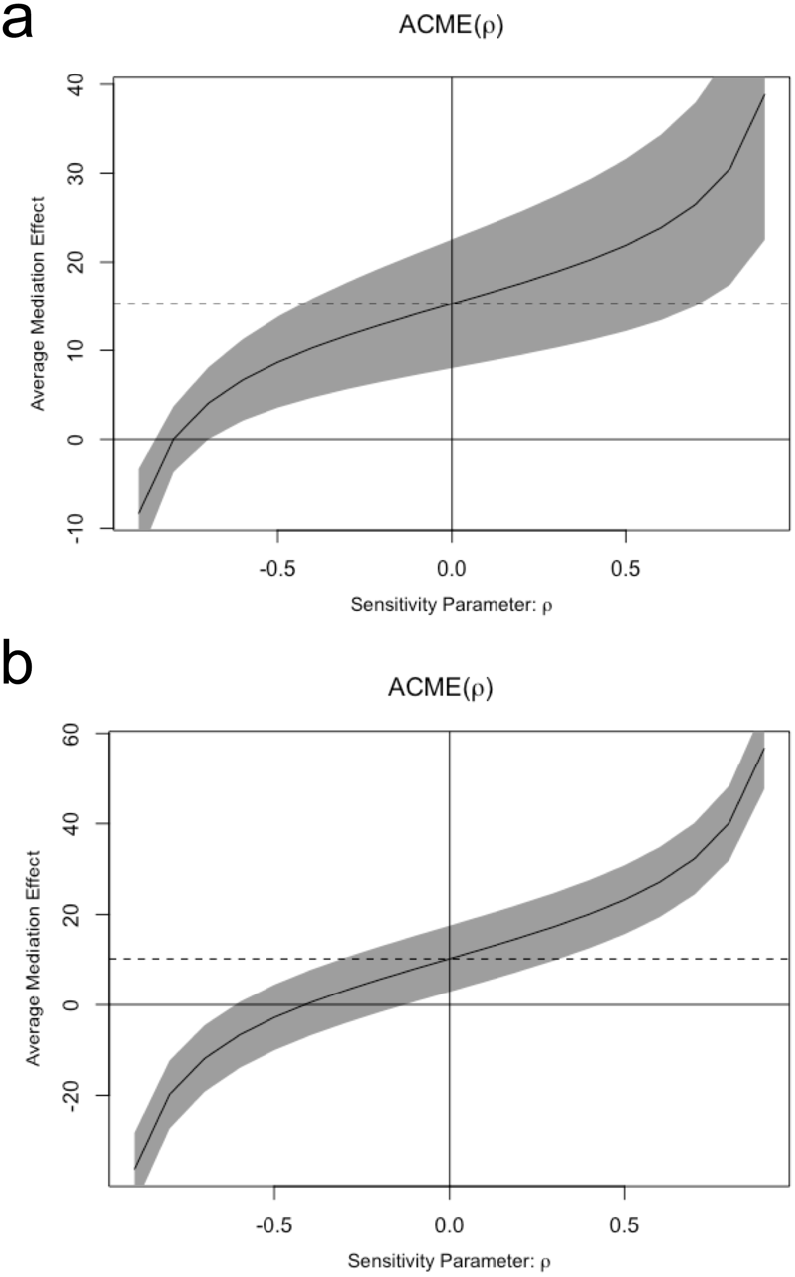
Sensitivity analyses for Pons FA mediation of cumulative blast exposure effects. **a)** Sensitivity analysis for mediated effects on TBI-associated neurobehavioral symptoms using NSI+ total score. The results show that for the point estimate of the to be zero, the correlation of ACME between the adjusted Pons FA value and PSQI total score must be approximately -0.9. b) Sensitivity analysis for mediated effects on sleep using PSQI total score. The results show that for the point estimate of the to be zero, the correlation of ACME between the adjusted Pons FA value and PSQI total score must be approximately -0.4. Gray area is the 95% CI.

